# In-person schooling and associated COVID-19 risk in the United States over Spring Semester 2021

**DOI:** 10.1101/2021.10.20.21265293

**Authors:** Kirsten E. Wiens, Claire P. Smith, Elena Badillo-Goicoechea, Kyra H. Grantz, M. Kate Grabowski, Andrew S. Azman, Elizabeth A. Stuart, Justin Lessler

## Abstract

Because of the importance of schools to childhood development, the relationship between in-person schooling and COVID-19 risk has been one of the most important questions of the COVID-19 pandemic. Previous work using data from the United States in winter 2020-21 showed that in-person schooling carried some risk for household members, and that mitigation measures reduced this risk. However, in-person schooling behavior and the COVID-19 landscape changed radically over the 2021 spring semester. Here we use data from a massive online survey to characterize changes in in-person schooling behavior and associated risks over that period. We find a significant increase in the frequency of in-person schooling and a reduction in mitigation, and that in-person schooling is associated with increased reporting of COVID-19 outcomes, even among vaccinated individuals (though the absolute risk among the vaccinated is greatly reduced). Moreover, vaccinated teachers working outside the home were less likely to report COVID-19-related outcomes than unvaccinated teachers reporting no work outside the home. Adequate mitigation measures appear to eliminate the excess risk associated with in person schooling.

## Introduction

The role of children and in-person schooling in SARS-CoV-2 transmission continues to be a contentious issue. Policies regarding in-person schooling have varied dramatically across school districts in the United States, with a heterogenous mix of in-person and remote learning, as well as varying approaches to mitigation (*1*). Over the spring semester of the 2020-21 school year many school districts made major updates to their approach to in-person schooling as the winter wave of the COVID-19 pandemic receded. Unfortunately, resurgences related to the Delta variant in the fall of 2021 have meant that, at the time of writing, COVID-19 remains a major health threat in the US and world-wide (*2, 3*). However, in light of increased vaccine availability and a broad consensus on the benefits of in-person schooling, the vast majority of school districts in the United States are conducting in-person classes despite this new surge in cases (*4*). Hence, understanding the risks associated with in-person schooling and how best to control them remains an important area of research.

A challenge in quantifying the risk of in-person schooling has been limited information on the degree to which children, who mostly have mild or asymptomatic infections, can infect teachers and family members (*5*). In an effort to address this issue, we previously analyzed data collected by the nation-wide US COVID-19 Trends and Impact Survey (US CTIS) between November 24, 2020 and February 10, 2021 (*6*). This survey is administered by the Delphi Group in partnership with Facebook and includes questions on demographics, symptoms related to COVID-19, positive SARS-CoV-2 test results, and schooling for any children in the household (*13,14*). We showed that individuals in a home with a child engaged in in-person schooling were at significantly higher risk of developing COVID-19-related outcomes (*7*). We also found that this risk decreased with increasing numbers of school-based mitigation measures, with no additional risk – as compared to no in-person schooling – associated with children attending schools with seven or more mitigation measures (*7*).

Between the time of this original study and the end of the 2021 spring semester in June, there were major changes in both the pandemic situation and schooling policies in most US schools. These include rising vaccination rates and the rise of the Alpha (and later the Delta) variant in the US, both of which had major impacts on the dynamics of the COVID-19 epidemic (*8*). The proportion of the US adult population who received at least one COVID-19 vaccine dose increased from about 7% at the end of January to 53% by mid-June (*9*). Beginning in mid-February 2021, the Alpha variant, which is roughly 50% more transmissible than the previously dominant SARS-CoV-2 strains (*10*), spread rapidly throughout the country (*3, 11*). In June, the Delta variant, which is about 60% more transmissible than Alpha (*12*), began to dominate (*3, 11*). It was unclear how these factors modified the risk posed by in-person schooling.

In this study, we expand our previous analysis (*7*) to include data from the US CTIS for the entire 2021 spring semester (which includes four weeks from the previous study), with the goals of 1) characterizing how rates of in-person schooling and implementation of school-based mitigation measures changed over the course of the semester, 2) understanding if and how vaccination status and variant prevalence modified the association between household COVID-19 risk and living with a child in in-person schooling, and 3) identifying other temporal trends in the relationship between in-person schooling and the risk of household members reporting COVID-19-related outcomes.

## Results

We analyzed data from 1,082,773 respondents living with school-aged children in 50 US states and Washington, DC that were collected through the Delphi Group at Carnegie Mellon University US COVID-19 Trends and Impact Survey (US CTIS) from January 12, 2021 to June 12, 2021 (Table S1). Though the total number of respondents decreased over the study period, patterns in the relative number of respondents by county remained consistent (Fig. 1a).

**Fig. 1.**
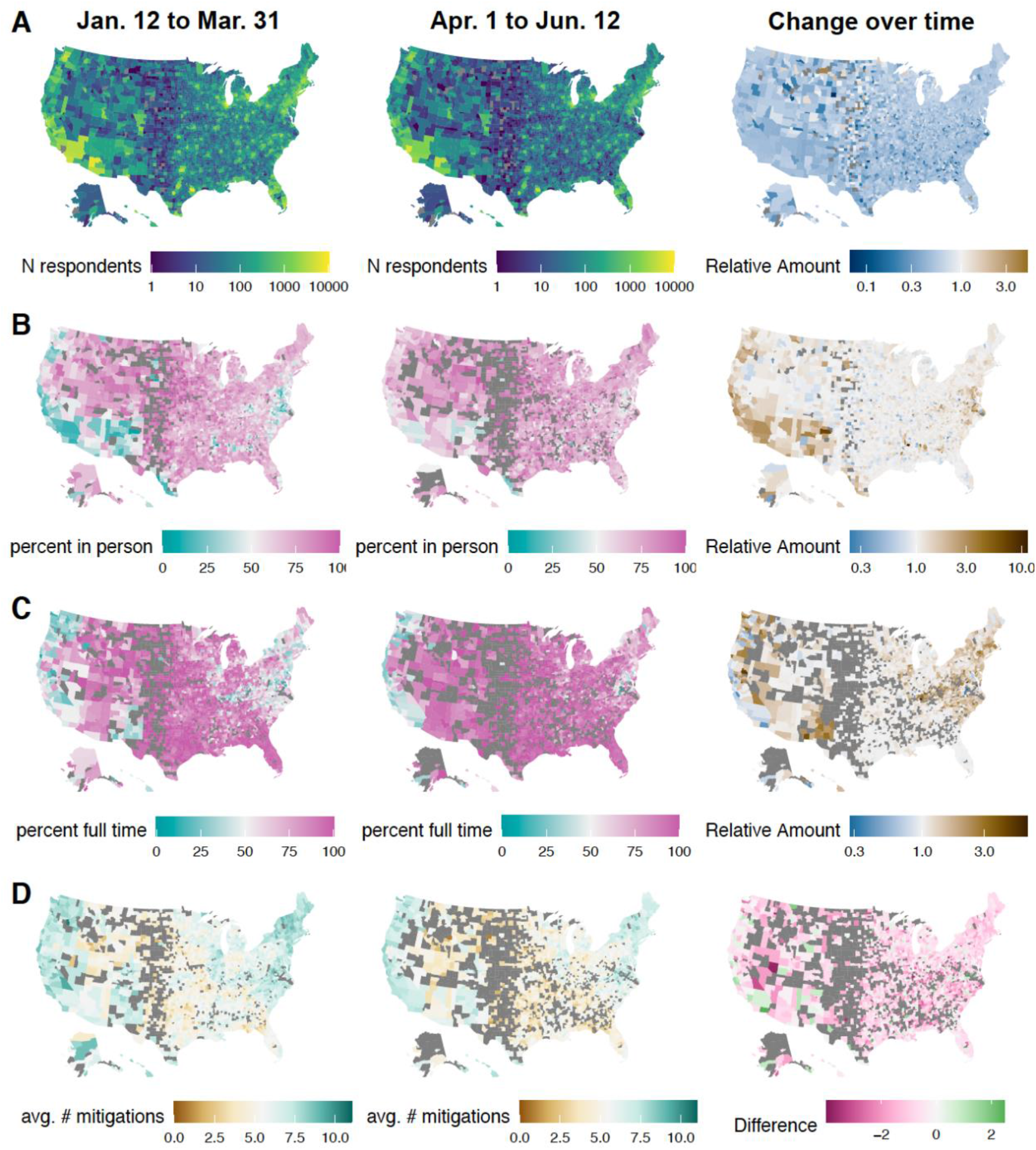
Changes over time in in-person schooling by county. Distribution of survey responses from Jan. 12 to Mar. 31 (left column), Apr. 1 to Jun. 12 (center column) and change over time (right column). Results are shown for (**A**) number of survey respondents reporting ≧1 school-aged child in the household, (**B**) percent reporting in-person schooling, (**C**) percent of respondents with in-person schooling reporting full-time in-person instruction, and (**D**) average number of school-based mitigation measures. “Relative Amount” in the right column indicates values in Apr. 1 to Jun. 12 (center column) divided by values in Jan. 12 to Mar. 31 (left column). Grey indicates county/periods where fewer than 10 respondents reported in-person schooling.

Overall, 59.4% of respondents living with school-aged children reported having at least one child in their household attending in-person schooling. The proportion of respondents living with school-aged children reporting any in-person schooling increased from 47.0% during the week of January 12, 2021 to 65.3% during the week ending June 12, 2021 (Fig. 1b, Fig. 2a). Of those reporting any in-person schooling, 74.0% reported full time in-person instruction, increasing from 69.0% to 82.1% over the study period (Fig. 1c).

**Fig. 2.**
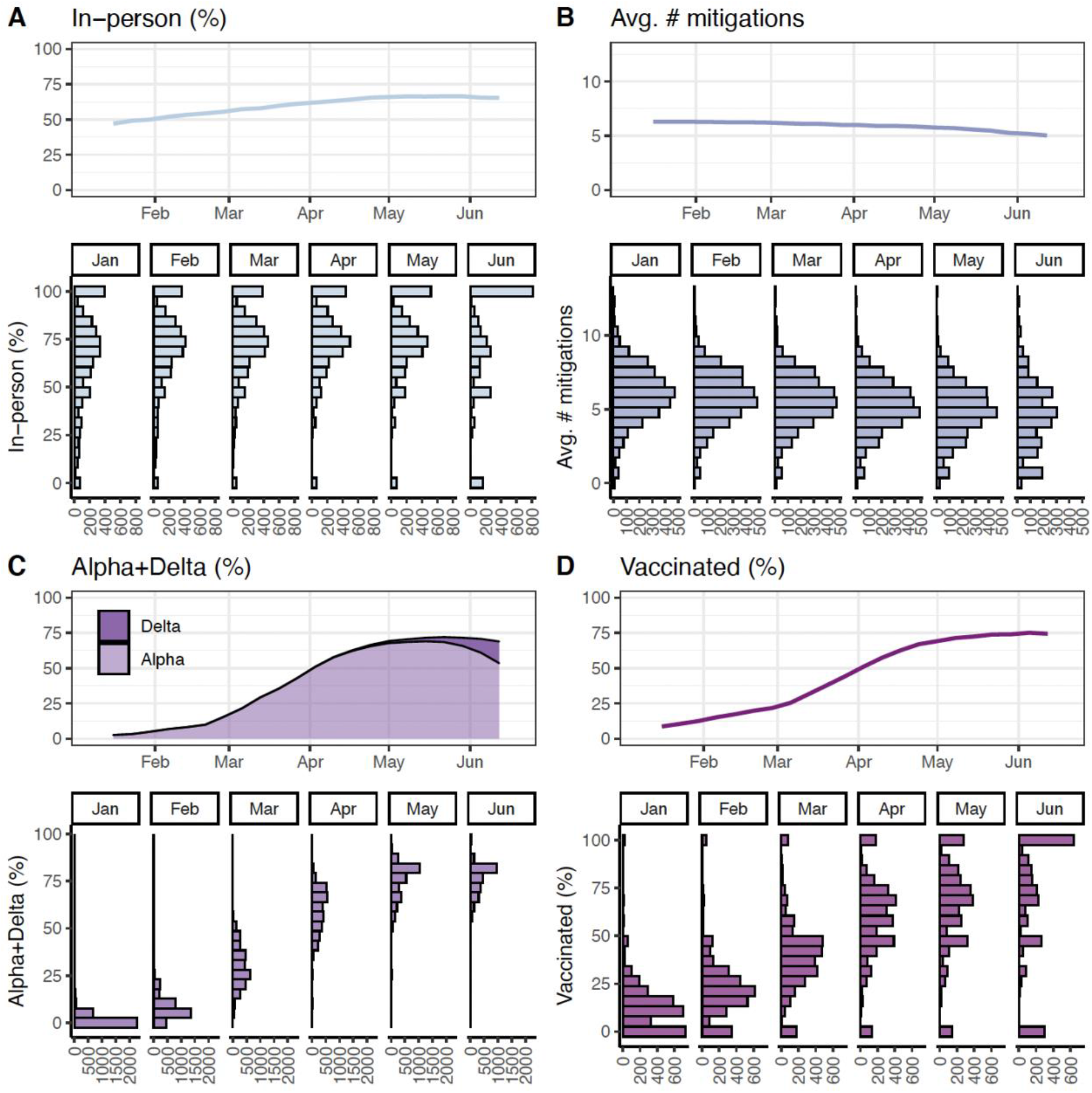
Concurrent changes in in-person schooling, vaccination, and variant prevalence. Changes over time between Jan. 12 and Jun. 12 in (**A**) percent of respondents living with school-aged children reporting any in-person schooling, (**B**) average number of school-based mitigation measures, (**C**) smoothed percent of GISAID SARS-CoV-2 sequenced isolates that were Alpha or Delta, and (**D**) percent of respondents living with school-aged children reporting having received at least one COVID-19 vaccine dose. Upper panel line plots show national averages by week weighted using US CTIS survey weights (**A-B**), state population (**C**), and county population (**D**). Lower panel histograms show the number of county-months with the indicated percentages (**A**,**C**,**D**) or numbers (**B**). Note that the number of respondents decreased over time (Fig 1d), which may contribute to the increasing number of zero values in the histograms in June.

Of those respondents living with a child attending school in-person, 93.1% reported at least one mitigation measure in place, 72.2% reported at least four, and 46.9% reported at least seven. The average number of mitigation measures decreased from 6.3 to 5.0 over the study period (Fig. 1d, Fig. 2b).

The biggest changes in in person schooling behavior occurred in places with the least in-person schooling at the start of the study period (Fig. 1b-1d), though the geographic regions with the most cautious approaches to in person instruction remained largely the same.

Since the 7th wave of the survey, starting on January 12, 2021, the US CTIS has included questions on the respondent’s vaccination status and number of doses received (*13, 14*). Overall, 43.1% of 1,082,773 respondents living with school-aged children reported having received at least one COVID-19 vaccine dose, compared with 50.8% among all 5,273,116 survey respondents. Though vaccination among those with school-aged children was lower on average (Fig. S1a), the magnitude of these differences varied by county (Fig. S1b). Vaccination among survey respondents living with school-aged children was strongly correlated with rates reported by the CDC (*15*) (Pearson’s r = 0.83), but rates were higher among survey respondents in most counties (particularly those in New Mexico, South Dakota, and Nebraska) (Fig. S1c, S1d).

While in-person schooling increased over the study period, vaccination rates and the prevalence of Alpha and Delta variants increased to a greater extent (Fig. 2, Movie S1). Because of the low proportion of cases from the Delta variant over the study period, we analyze the Alpha and Delta variants together throughout this manuscript. The prevalence of Alpha/Delta variants circulating in the population increased from 1.7% in the week of January 12 to 72.6% in the week ending June 12th (Fig. 2c, Movie S1). At the same time, the percentage of respondents living with school-aged children that reported having received any number of COVID-19 vaccine doses increased from 8.7% to 74.4% (Fig. 2d, Movie S1). Monthly rates of in-person schooling, vaccination, and average number of mitigation measures varied widely across counties, and this variation persisted throughout the spring semester (Fig. 2a,b,d).

The percent of respondents living with school-aged children that reported COVID-19-like illness (CLI; defined as fever of at least 100°F, as well as cough, shortness of breath, or difficulty breathing) remained similar across the study period, ranging from 1.83% in January to 1.99% in June (Table S2). The more specific indicators of potential SARS-CoV-2 infection, loss of taste and/or smell and a positive SARS-CoV-2 test with the past 14 days, decreased from 3.65% to 2.17% and 3.44% to 0.09% respectively (Table S2). These temporal trends were similar among all survey respondents and among respondents stratified by in-person schooling status; though, respondents living with school-aged children not participating in any in-person schooling saw a decrease in CLI over the study period, from 1.36% to 0.97% (Table S2).

Overall—from January 12 to June 12, 2021—after adjusting for county-level SARS-CoV-2 biweekly attack rates averaged over the past four weeks, COVID-19 vaccination status, and other individual- and county-level factors, living in a household with a child in full-time in-person schooling was associated with increased odds of CLI (aOR, 1.32; 95% CI, 1.25 to 1.40), losing taste and/or smell (aOR, 1.19; 95% CI, 1.15 to 1.24), and reporting a positive SARS-CoV-2 test (aOR, 1.32; 95% CI, 1.27 to 1.38) (Fig. 3). In contrast to our previous analysis (*7*), we saw no clear trends by grade in any of the COVID-19-related outcomes associated with in-person schooling (Fig. 3, Fig. S2).

**Fig. 3.**
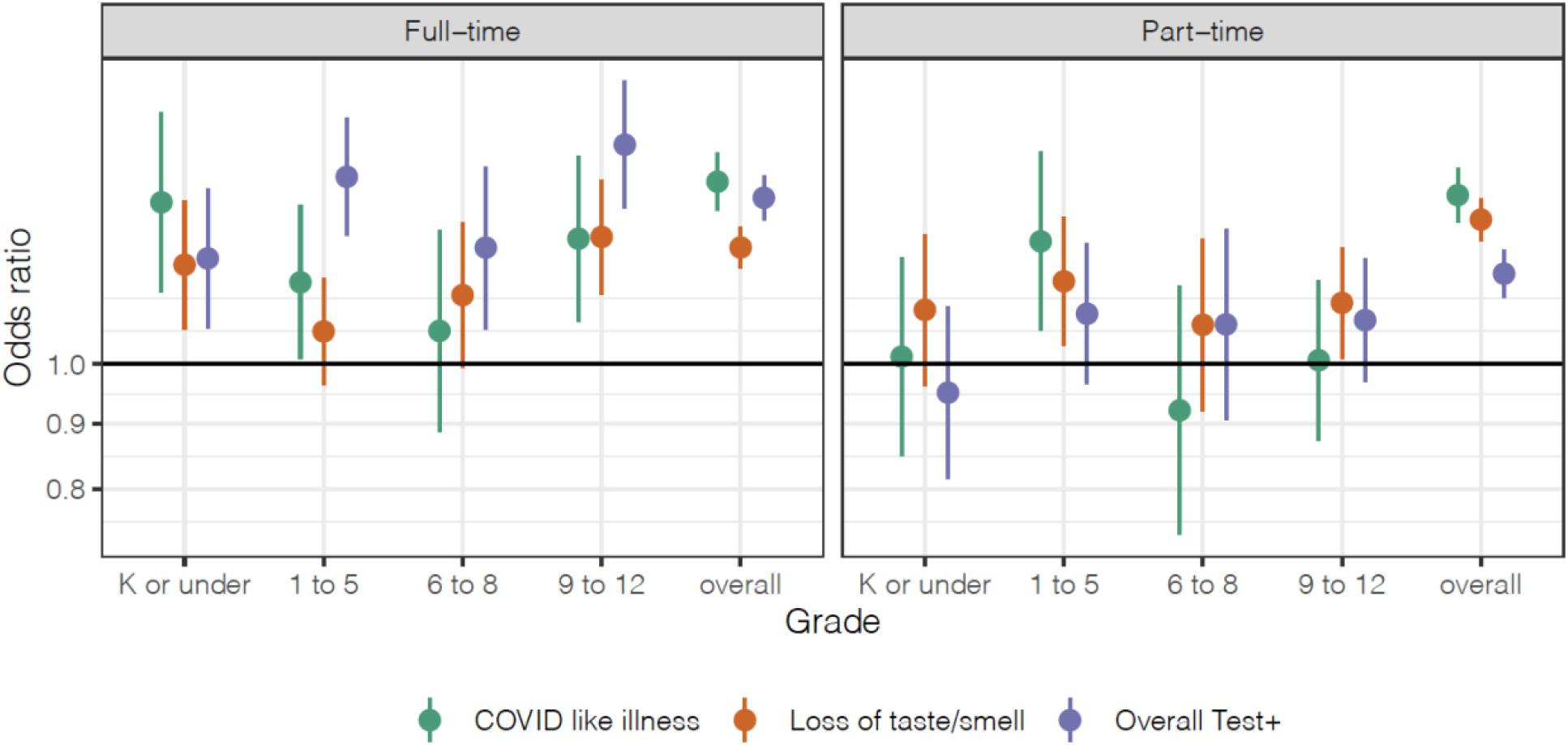
Risk associated with in-person schooling overall and by grade level. Odds ratios of COVID-19-related outcomes by full- and part-time in-person schooling compared to no in-person schooling, overall and stratified by grade, adjusted for individual- and county-level covariates across the study period from Jan. 12 to Jun. 12. **Note:** grade-stratified analyses include only those respondents who reported living with school-aged children in a single grade category, while the overall analyses include all respondents living with school-aged children; thus, the overall estimates do not always fall in-between the grade-specific estimates.

Consistent with our previous study (*7*), the most commonly reported mitigation measure was student masking, reported by 88% of respondents with in-person schooling in January and 84% in June (Fig. 4a). The second most common measure was teacher masking, at 76% in January and 62% in June (Fig. 4a). Among those with in-person schooling, teacher masking was associated with the greatest risk reduction across all COVID-19-related outcomes, followed by daily symptom screens, student masking and restricted entry (Fig. 4b). These trends were consistent over time, with larger uncertainty in risk estimates in May-June (Fig. S3). These patterns were also consistent for other households and individual level COVID-19-related outcomes (Fig. S4).

**Fig. 4.**
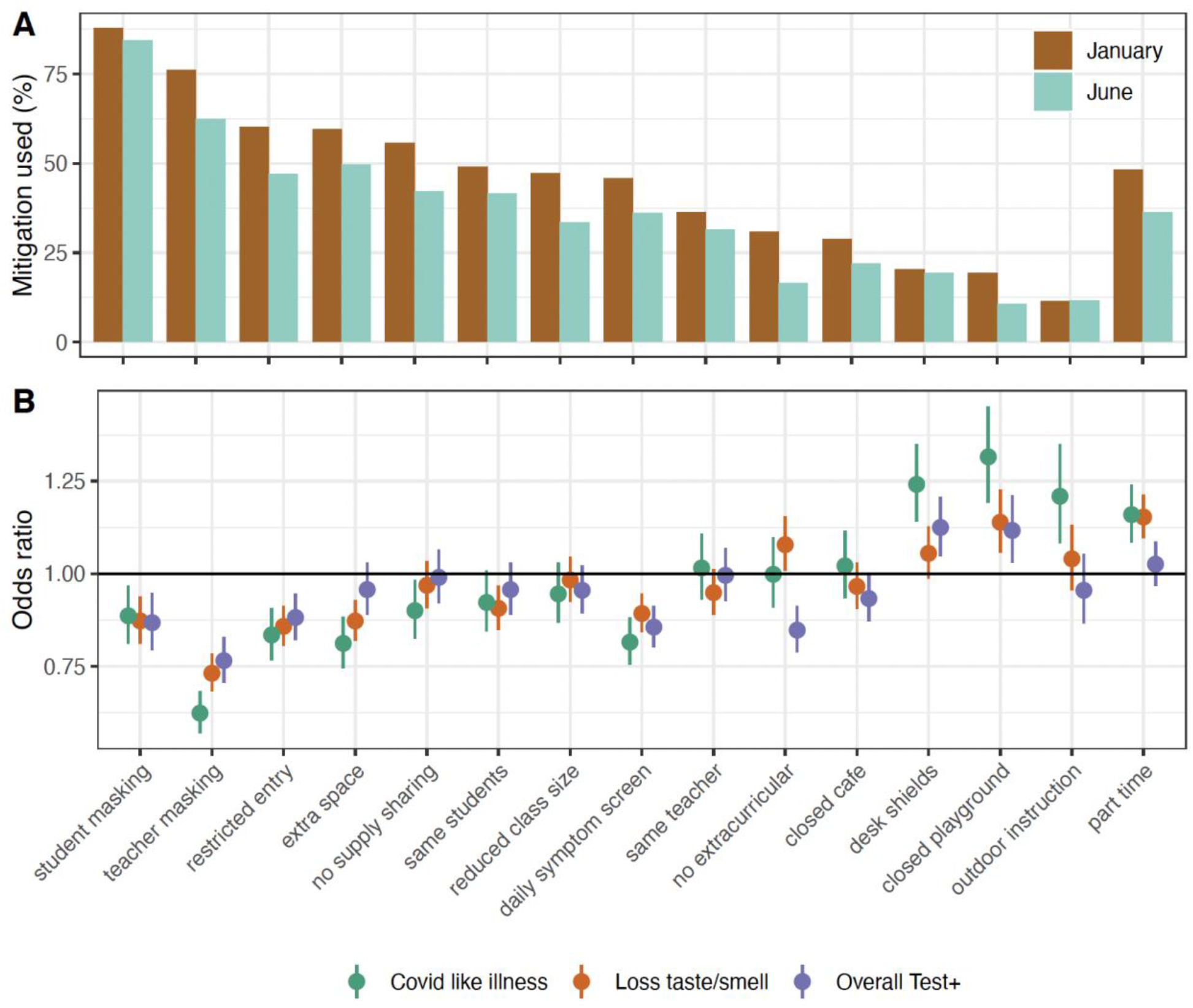
Individual mitigation measures. (**A**) Percent of respondents with in-person schooling that reported each mitigation measure being used in January and June, weighted using US CTIS survey weights. (**B**) Odds ratio of COVID-19-related outcomes among respondents with children in in-person schooling who reported each mitigation measure compared to those with children in in-person schooling who did not report each measure, adjusted for individual- and county-level covariates across the study period from Jan. 12 to Jun. 12.

Across the 2021 spring semester, the association between of CLI and loss of taste/smell and full-time and part-time in-person schooling disappeared with report of four or more school-based mitigation measures, and risk of a SARS-CoV-2 positive test disappeared with report of seven or more mitigation measures (Fig. 5). Results were consistent for other household and individual level COVID-19-related outcomes (Fig. S5). These patterns were consistent from January to February and March to April (Fig. 5). By May to June, risks of all COVID-19-related outcomes disappeared when four or more mitigation measures were reported (Fig. 5).

**Fig. 5.**
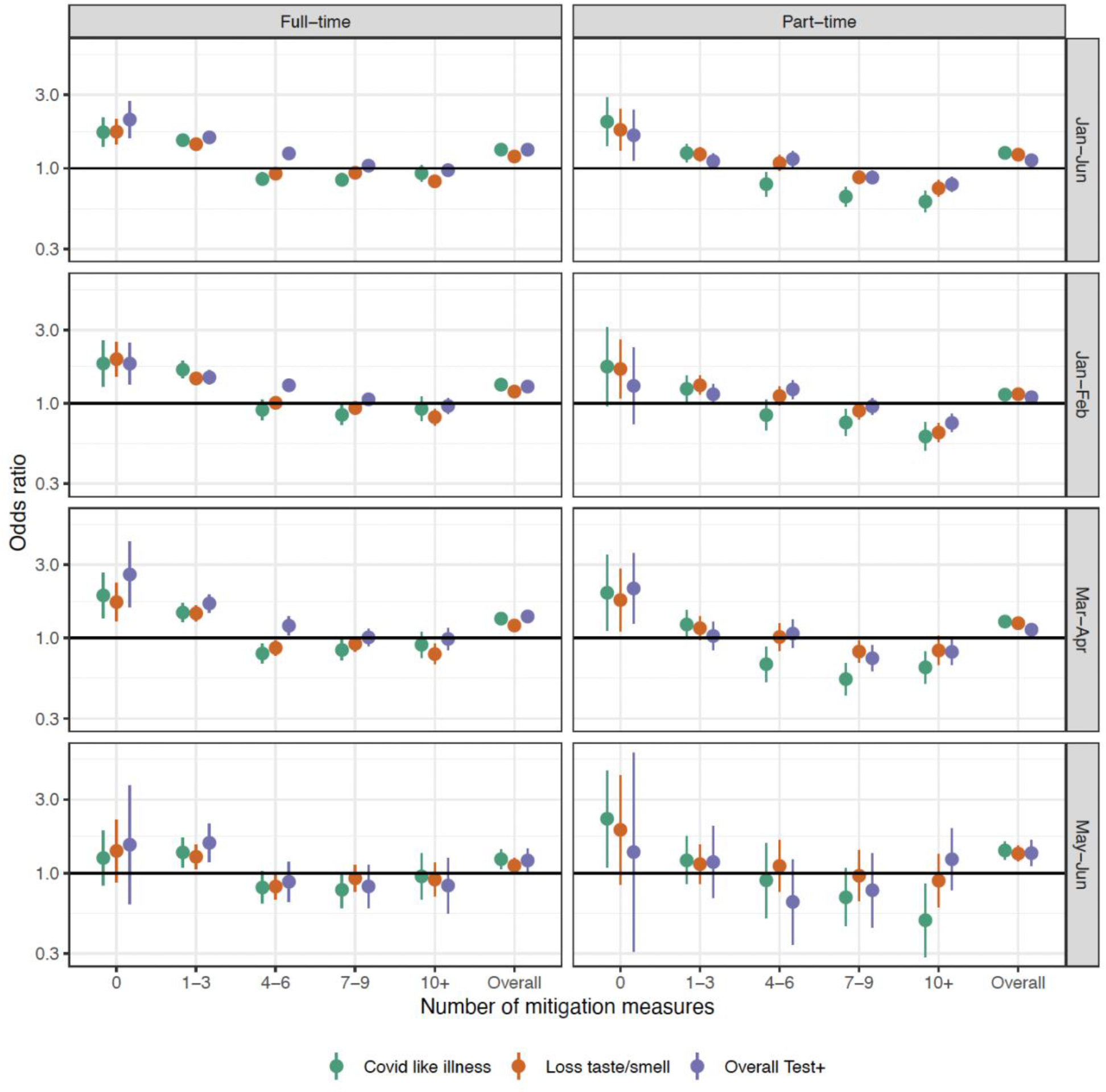
Risk associated with in-person schooling over time by number of reported mitigation measures. Odds ratio of COVID-19-related outcomes by full- and part-time in-person schooling and number of school-based mitigation measures compared to no in-person schooling, adjusted for individual- and county-level covariates. “Overall” indicates adjusted odds ratios of full-time and part-time schooling with any number of mitigation measures compared to no in-person schooling. Odds ratios are shown across the entire study period from Jan. 12 to Jun. 12. (Jan-Jun), as well as within time periods from Jan.12 to Feb. 28 (Jan-Feb), Mar. 1 to Apr. 30 (Mar-Apr), and May 1 to Jun. 12 (May-Jun).

Over the study period, each ten percent increase in the state-level prevalence of Alpha/Delta variants was associated with increased baseline risk of CLI (aOR 1.05; 95% CI, 1.03 to 1.07), loss of taste and/or smell (aOR 1.02; 95% CI, 1.01 to 1.03), and reporting a positive SARS-CoV-2 test (aOR 1.05; 95% CI, 1.03 to 1.06), after adjusting for background incidence, vaccination status, and other individual- and county-level characteristics. The rise of Alpha/Delta variants did not change the relative association between in-person schooling and COVID-19-related outcomes (Fig 6). These findings did not change noticeably when we additionally adjusted for cumulative incidence of confirmed SARS-CoV-2 as an indirect indicator of population immunity (Fig. S6).

**Fig. 6.**
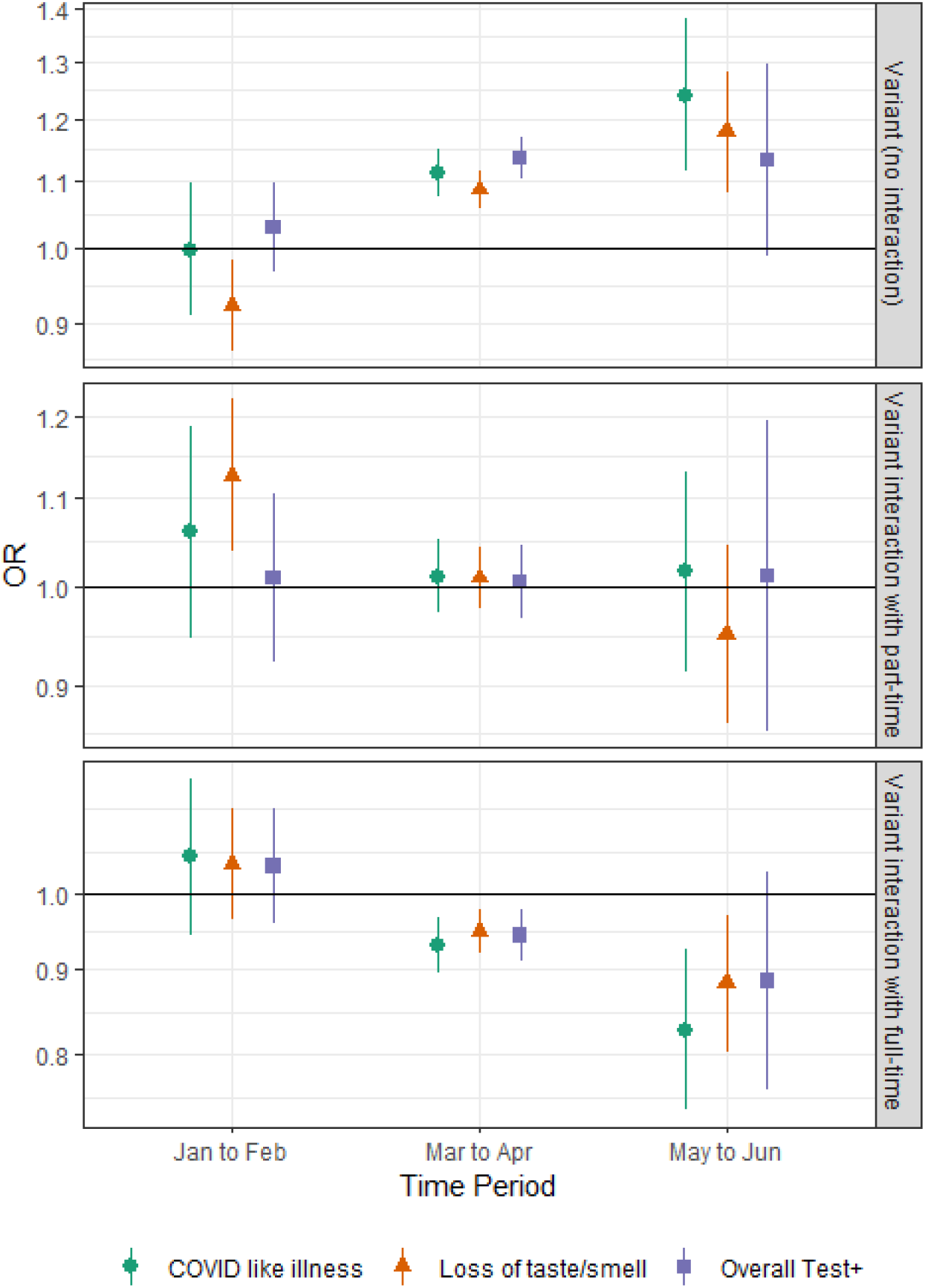
Risk associated with in-person schooling and Alpha/Delta variants over time. Estimated baseline risk (top panel) associated with the proportion of cases due to the Alpha and Delta variants and interaction with part-time (middle panel) and full-time (bottom panel) in-person schooling status. Interaction terms show the additional impact of the variant on top of baseline risk due to part- and full-time in-person schooling. Risk is shown within two-month time strata.

We next examined if and how the relationship between respondent vaccination status and living in a household with a child in in-person schooling varied over time (Fig. 7). We found that, by the end of the study period, there was a modest correlation between increased in-person schooling and lower vaccination rates at the county level (Pearson’s r = −0.15; 95% CI, −0.18 to −0.12), while there was a positive correlation between vaccination rates and the number of school-based mitigation measures reported in a county (Pearson’s r = 0.36; 95% CI, 0.32 to 0.39).

**Fig. 7.**
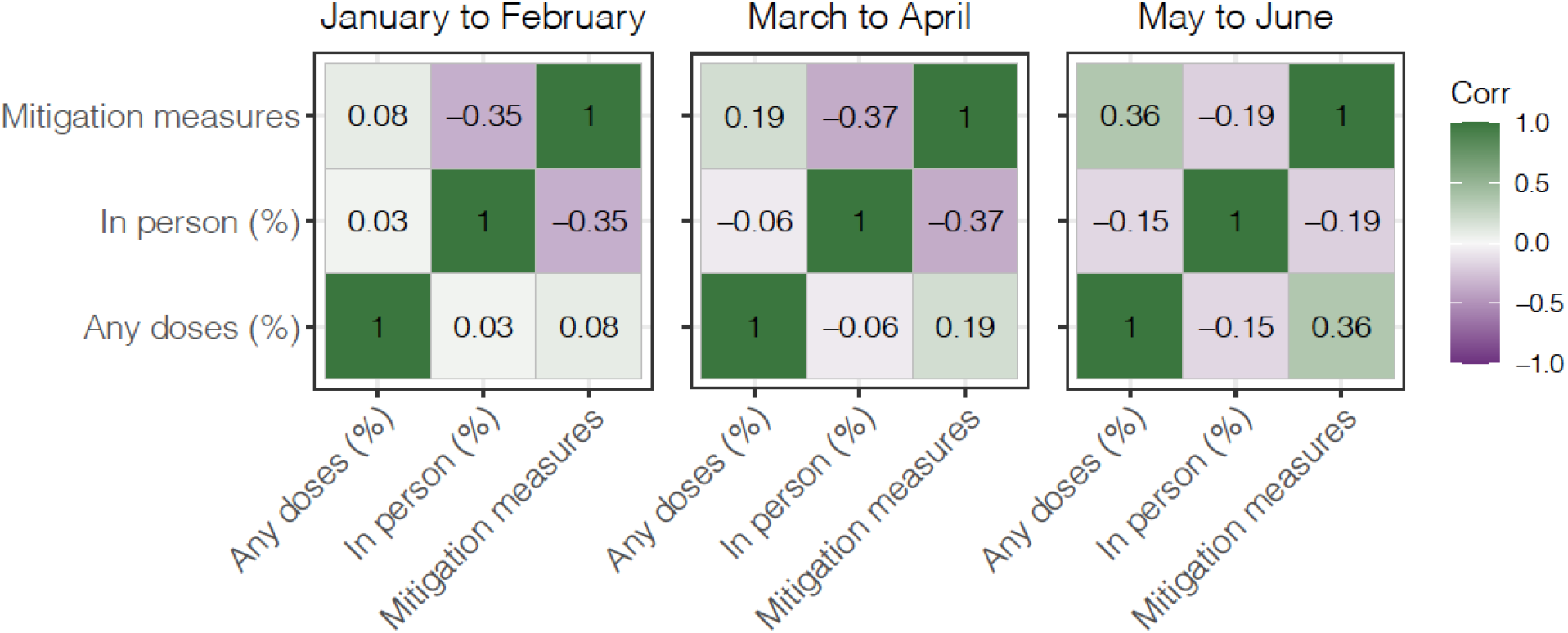
Correlations between vaccination, in-person schooling, and mitigation measures. Pearson’s correlation coefficients between percent vaccinated with at least one dose of a COVID-19 vaccine, percent of respondents living with school-aged children reporting in-person schooling, and average number of school-based mitigation measures by county-month. Results are shown within time periods Jan. 12 to Feb. 28 (left), Mar. 1 to Apr. 30 (center), and May 1 to Jun. 12 (right). All coefficients shown were significant (p < 0.05). Limited to county-months with at least 5 respondents.

We explored the combined impact of living with a child in in-person schooling and vaccination status on COVID-19-related outcomes, and whether interactions existed between the two risk factors (Fig. 8; Table S7). Overall, we found a reduction in the odds of COVID-19-related outcomes associated with both no in-person schooling and vaccination, with the latter having the far larger impact (Fig. 8a). Those having received two vaccine doses and not engaged in in-person schooling had the lowest risk of reporting COVID-19-related outcomes by a large margin (Fig. 8a). In-person schooling did not modify the association between vaccination status and reporting CLI but was, unexpectedly, associated with some variation in the apparent impact of vaccination on loss of taste/smell and reporting a positive SARS-CoV-2 test (Fig. 8b). The relative increase in the odds of reporting COVID-19-related outcomes associated with in-person schooling was the same regardless of vaccination status (Fig. 8c).

**Fig. 8.**
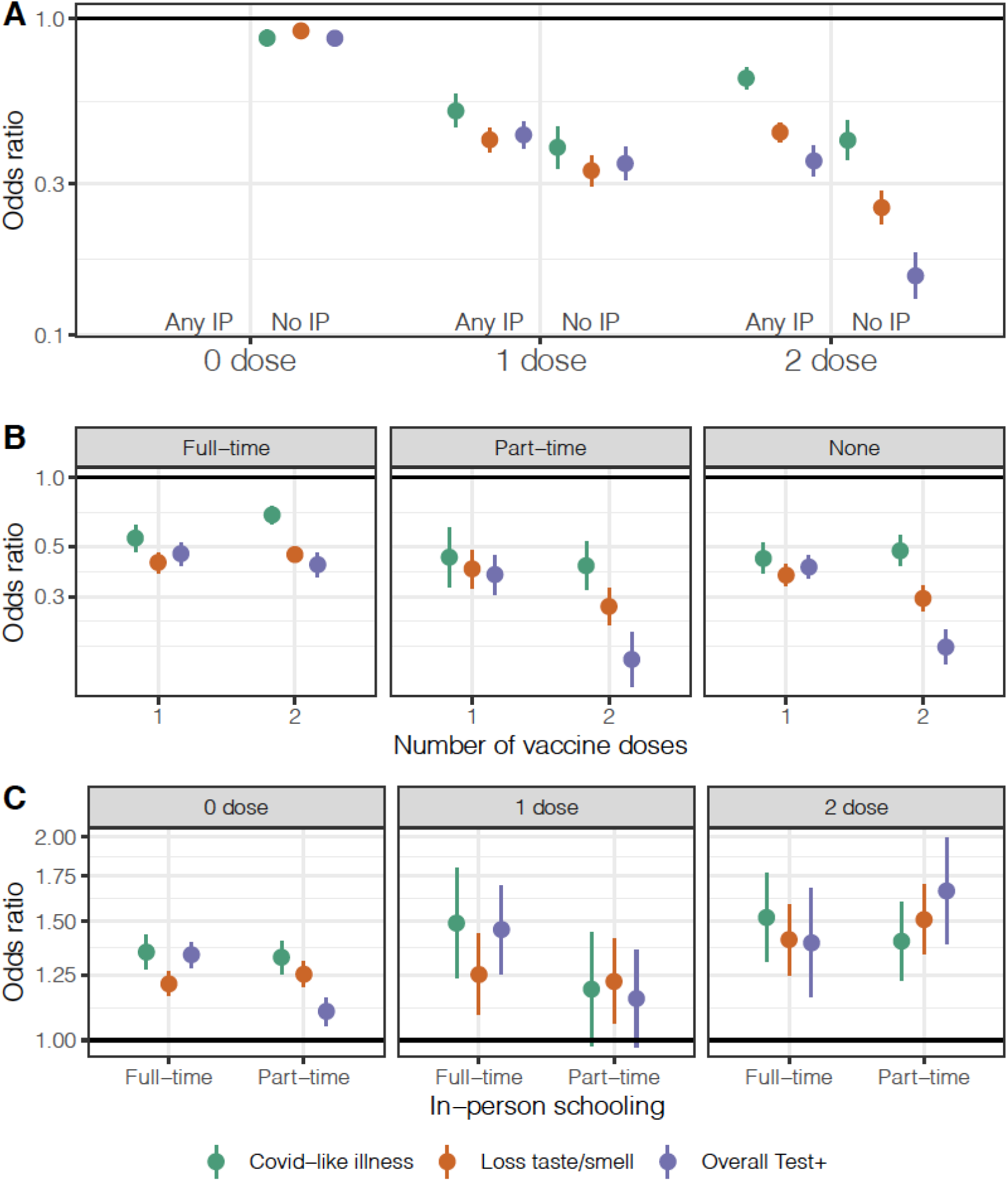
Risk associated with in-person schooling and number of vaccine doses. (**A**) Odds ratio of COVID-19-related outcomes by number of reported vaccine doses (0, 1 or 2) and in-person schooling status (living in a household with a child participating in any in-person (Any IP) or no in-person (No IP) schooling), adjusted for individual- and county-level covariates. Zero vaccine doses and any in-person schooling is the reference group. (**B**) Odds ratio of COVID-19-related outcomes with one and two vaccine doses compared to zero doses, when data are stratified by none, part-time, and full-time in-person schooling. (**C**) Odds ratio of COVID-19-related outcomes with part- and full-time in-person schooling compared to no in-person schooling when data are stratified by vaccine doses.

To understand COVID-19-related risk among teachers, we analyzed data from the 116,014 K-12 teachers included in the full US CTIS survey, whether or not they lived with a child participating in in-person schooling. We found that 86.0% of K-12 teachers reported work for pay conducted outside of their home in the previous four weeks. The percentage of teachers reporting paid work outside of the home increased from 77.5% in January to 92.4% in June, which was the largest increase in work outside the home for any occupation group (Fig. S7). In comparison, the proportion of office and administrative support professionals that reported working outside of the home increased by a much smaller amount, from 61.5% to 66.9% (Fig. S7).

Overall, being a K-12 teacher conducting any work outside the home was associated with higher risk of losing taste/smell (aOR 1.37; 95% CI, 1.13 to 1.65) and receiving a positive SARS-CoV-2 test (aOR 2.04; 95% CI, 1.67 to 2.48) compared to K-12 teachers working exclusively from home (Fig. 9). However, we found no differences in the risk of reporting COVID-19-related outcomes between teachers and office and administrative support professionals working outside the home (other professions also had similar risk). These trends held across time periods (Fig. S8; Table S8). Notably, vaccinated teachers working outside the home were less likely to report COVID-19-related outcomes than unvaccinated teachers reporting no work outside the home (Fig. S9).

**Fig. 9.**
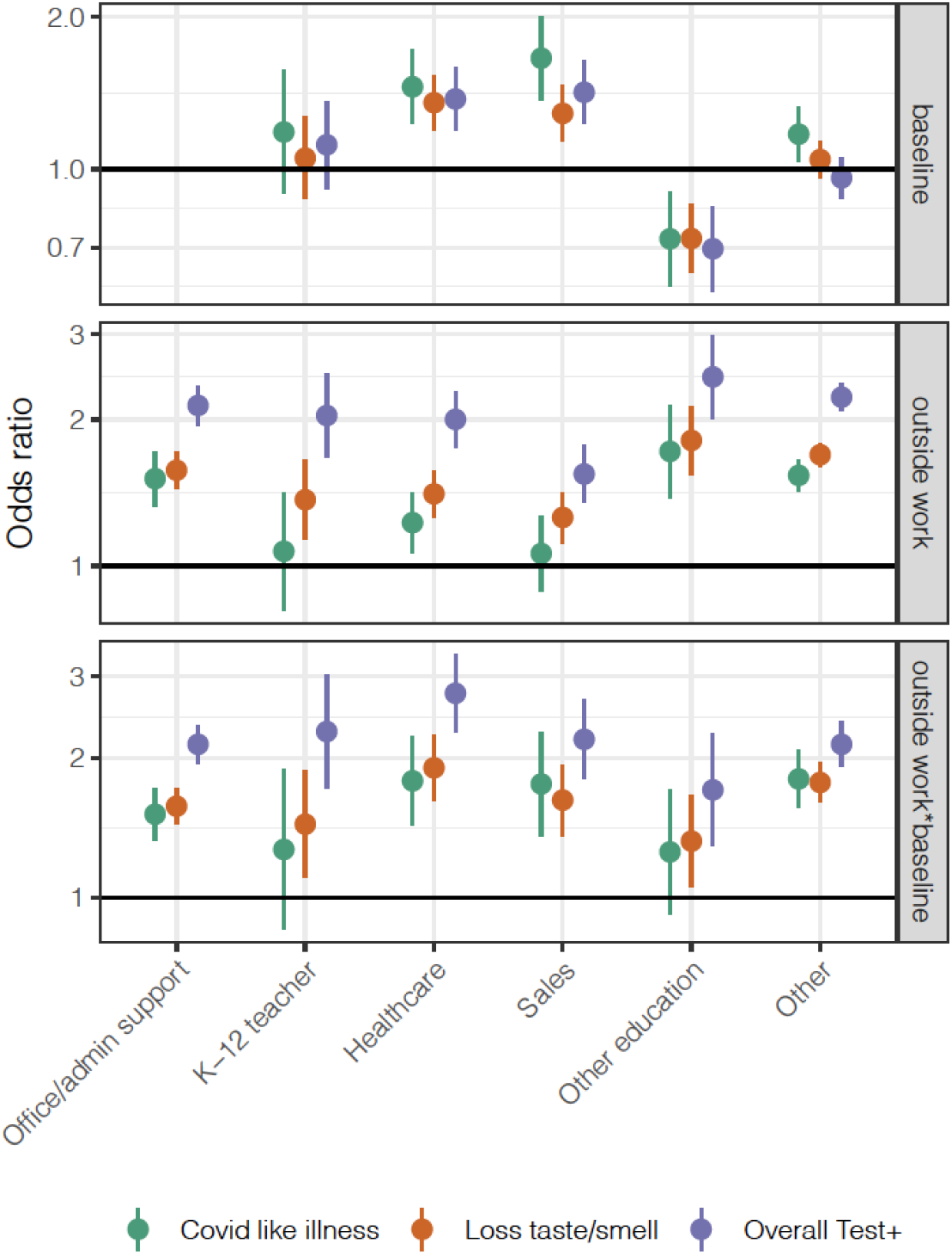
Risk by occupation and paid work outside the home. Odds ratio of COVID-19-related outcomes, contrasting office workers not reporting extra-household work for pay to those in other employment categories not reporting work for pay outside the home (top), and to those reporting work for pay outside the home (bottom). The middle row shows the odds ratio (i.e., increased risk) within each category associated with working outside the home compared to no work outside the home. Food service workers are excluded from this analysis because less than 5% reported working exclusively from home (Fig. S7).

## Discussion

In-person schooling increased across the US over the spring semester 2021, a period which also saw increasing COVID-19 vaccination rates and the spread of SARS-CoV-2 variants. In this study we show that, despite these changes, associations seen in winter 2020-21 (*7*) hold: in-person schooling is associated with increased risk of COVID-19-related outcomes in the household, but this risk can be reduced or eliminated with implementation of multiple mitigation measures in schools (Fig. 5, Fig. S5). These findings were consistent across the study period even as vaccination rates increased, emphasizing the importance of layered mitigation measures to reduce the risk of transmission in schools. Such measures remain crucial in light of increased COVID-19 cases due to the Delta variant (*3*) and the potential for future outbreaks, and are consistent with CDC guidance (*16*).

We also found that changes in in-person schooling and mitigation measures varied between states and counties, which has likely contributed to heterogeneity in risk. We found that higher levels of in-person schooling were accompanied by lower levels of mitigation measures (Figs. 1,2,7). We also found that from March to April, and even more so from May to June, vaccination rates were positively correlated with mitigation measures and inversely correlated with in-person schooling (Fig. 7). This tendency of communities to eschew both kinds of control may lead to significant increases in overall COVID-19 risk in these areas.

We found that being unvaccinated and living with a student engaged in in-person schooling was associated with the highest risk of reporting COVID-19-related outcomes, while, unsurprisingly, those reporting two doses of vaccine and no one in the household engaged in in-person schooling had the lowest risk (Fig. 8a). Even among individuals with two vaccine doses, in-person schooling was associated with significantly increased risk of COVID-19-related outcomes in household members compared to individuals with no children in in-person schooling (Fig. 8c). In other words, our results suggest that even though adult vaccination substantially reduces the overall risk from living with a child engaged in in-person schooling, the relative change in risk due to in-person schooling is similar to that seen in the unvaccinated.

The proportion of teachers working outside the home increased more than any other professional group over the spring semester 2021 (Fig. S7). Consistent with the in-person schooling results, K-12 teachers working for pay outside the home were at increased risk of COVID-19-related outcomes (Fig. 9), although the additional risk was similar to that in other occupations (Fig. 9). Importantly, vaccinated K-12 teachers working outside the home reported fewer COVID-19-related outcomes than the unvaccinated not working outside their homes. This emphasizes the critical role that vaccination can play in a safe return to the classroom for teachers.

Furthermore, similar to previous work (*7*), we found that teacher masking, student masking, restricted entry, and symptom screens were individually associated with the greatest reduction in the risk associated with in-person schooling (Fig. 4). We also found a particularly strong signal for CLI for teacher masking, which may indicate that masks are preventing the spread of other respiratory infections. In addition, we found that some of the less commonly used mitigation measures, such as desk shields and closed playgrounds, were associated with increased risk (Fig. 4). This may reflect reduced utility of these measures and/or a saturation effect since these are often in place alongside other mitigation approaches. Closed extracurricular activities were associated with reduced risk of positive tests in both respondents and family members, but not CLI or loss of taste/smell (Fig. 4, Fig. S4), which could reflect increased testing in households with students in extracurricular activities.

Although our results do not indicate that the rise of the Alpha variant over the study period changed the risk associated with in-person schooling, there were some unexpected patterns in the interaction between full-time schooling and variant prevalence, which may reflect a limitation of the data. As the prevalence of the Alpha variant was increasing in all states over the study period, as well as within each time strata we investigated, some of this effect may be due to other time-varying factors that are not captured in the data such as changing compliance with intervention measures. Additionally, data on variant prevalence was only available at the state-level, which may obscure important county-level differences.

This study has several additional limitations. The survey data that informed the risk estimates were self-reported and subject to recall bias. They were also gathered through the Facebook platform and may not be representative of the underlying populations, though this should be accounted for at least in part through the survey weights (*19*). We found that vaccination rates were higher in the survey data compared to CDC’s reported data (*15*) (Fig. S1). While CDC’s reported vaccination data is known to be under-reported, this could also reflect a bias in the survey data (*20*). For example, the US CTIS survey may be capturing a more affluent, less rural population, which could explain in part why we found particularly large differences in the survey data compared to CDC data in states with large rural populations (Fig. S1b).

We were also limited by data available to us in the survey or at the county-level. In our analysis of risk by occupation, we were unable to examine risk by the amount of time spent working outside of the home. In addition, there may be confounding factors that we were not able to adjust for, such as community-level vaccination and immunity. Though, additionally adjusting for cumulative incidence as a proxy for this did not noticeably impact on our results (Fig. S6). We were also not able to evaluate timing of vaccine doses in relation to COVID-19-related outcomes. Thus, it is possible that some of the individuals who received two vaccine doses had been infected prior to developing immunity. Finally, out of the seven COVID-related outcomes we measured, none specifically assessed asymptomatic infection. In addition, SARS-CoV-2 test positivity requires seeking out a test, and CLI is not specific for COVID-19. That our findings were largely concordant between these varied outcomes supports our overall conclusions. Some of the differences we did find between CLI and SARS-CoV-2 positive tests could reflect increases in non-SARS-CoV-2 respiratory infections over the spring semester, as was found in a study in Hong Kong (*21*).

Overall, our findings support previous studies that have shown secondary transmission and outbreaks associated with in-person schooling and child care (*22–26*). While there are other studies that have shown relatively low risk of transmission in schools (*27–31*), SARS-CoV-2 mitigation measures were in place in each of these settings. Thus, there is abundant evidence to indicate that in-person schooling plays a role in SARS-CoV-2 transmission, but that this risk can be mitigated. Despite major changes in in-person schooling behavior and the epidemiological situation over the course of the spring semester 2021, the apparent relative risks associated with in person schooling – and the measures that worked to mitigate these risks – remained mostly the same. Hence, as we confront the current and possible future COVID-19 epidemics, these results can help guide us as we strive to keep our homes safe while not sacrificing the education of our children.

## Materials and Methods

### Survey data

We analyzed survey data collected by the US COVID-19 Trends and Impact Survey (US CTIS). The US CTIS is administered by the Delphi Group in partnership with Facebook and includes questions on demographics, symptoms related to COVID-19, positive SARS-CoV-2 test results, vaccination, and schooling for any children in the household (*13*); detailed survey questionnaires are available at (*14*). The survey employs two-stage sampling (*19, 32*), and throughout this study we used the survey weights to adjust for 1) differences between the US population and US Facebook users, and 2) the propensity of a Facebook user to take the survey. Our study period encompasses Waves 7-8 and 10-11 of this survey (*33*), covering the spring semester of the 2020-21 school year from Jan. 1 to June 12, 2021. Additional details can be found in the Supplementary Methods.

### Variant Data

Daily state-level SARS-CoV-2 variant data were obtained from outbreak.info (*35*), which utilizes data from GISAID (*11*). This data includes the daily proportion of sequences for each variant.For each state, a smooth spline with six degrees of freedom was fitted to the combined Alpha and Delta variant proportion over time. Each survey response was assigned the fitted variant proportion for their state corresponding to seven days before the survey response date.

### CDC Vaccine Data

Daily county-level vaccination data were obtained from the US Centers for Disease Control and Prevention (*15*) on July 14th, 2021. Weekly proportions of individuals in each county that had received at least one vaccine dose were calculated by taking into account county populations and percent completeness of the reported CDC data. These data were used in linear regression and Pearson’s correlation analyses to examine how county-level vaccination rates reported by the CDC compared with those reported by the Facebook survey respondents living in a household with school-aged children. The latter survey data were used as individual-level covariates to adjust for respondent vaccination status in the analyses described below and in the Supplementary Methods.

### COVID-19 Case Data

County-level COVID-19 case data were obtained through the Johns Hopkins Center for Systems Science and Engineering (JHU-CSSE) COVID-19 Dashboard (*34*). County-week-level attack rates were calculated as the average 2-week incidence in the past 4 weeks. Attack rates were calculated by dividing reported COVID-19 cases by 2020 county populations, which were obtained from the US Census Bureau using the tidycensus package (https://cran.r-project.org/package=tidycensus). Adjustment for attack rates was done based on the log base 2 cases per thousand (i.e., log2([average biweekly attack rate per 1000] + 1)).

### Covariates and Outcomes

In addition to average 2-week incidence in the past 4 weeks, county-level covariates (obtained from the 2014-2018 American Community Survey using the tidycensus package) included total population, percent of the population that was white, percent of households with income below the poverty threshold, a measure of income inequality, and metropolitan type. Individual- and household-level covariates (obtained from the US CTIS dataset) included gender, age, occupation, educational level, household size, masking behavior, out-of-state travel, vaccination with any number of COVID-19 vaccine doses, and whether they reported a visit to a bar/restaurant/cafe, to an event with more than 10 people, and whether they used public transit. Primary outcomes included COVID-19 like illness, loss of taste and/or smell, and a positive SARS-CoV-2 test results in the past 24 hours. Secondary outcomes included CLI in any household member, contact with a household member who received a positive test result, and a positive test result when the test was not indicated. Detailed descriptions of all variables and outcomes can be found in the Supplementary Methods.

### Analysis

All analyses were conducted using quasibinomial regression accounting for survey weights using the srvyr package (https://cran.r-project.org/package=srvyr) in the R statistical language. The overall analysis included adjustments for the baseline covariates described above as well as full- and part-time in-person schooling status of children living in the household. The analysis of COVID-19 risk and number of school-based mitigation measures included full- and part-time in-person schooling variables categorized by number of mitigation measures. The analysis of individual control measures was restricted to respondents living with children in in-person schooling and adjusted for baseline covariates as well as each individual school-based mitigation measure. The analysis of in-person schooling and Alpha/Delta variants included the baseline covariates, the fitted combined Alpha/Delta variant proportion, and two interaction terms between variant proportion and each of full- and part-time in-person schooling. The analyses of in-person schooling and number of vaccine doses included the baseline covariates, but with vaccination status replaced by the number of COVID-19 vaccine doses. The analysis of the risk of COVID-19-related outcomes among educational professionals was conducted among all respondents, regardless of living with a child in in-person schooling, and included baseline covariates as well as an interaction term between occupation types and an indicator for any paid work outside the home in the last four weeks. Full details of each analysis including regression formulas can be found in the Supplementary Methods.

## Data Availability

All data produced in the present work are contained in the manuscript

## Funding

This work was supported by the Johns Hopkins University Discovery Award (to E.B.G. and E.A.S.), the Johns Hopkins University COVID-19 Modeling and Policy Hub Award (to E.B.G. and E.A.S.), and the Department of Health and Human Services (to J.L. and M.K.G.).

## Author contributions

Conceptualization: J.L., M.K.G., A.S.A., and E.A.S. Methodology: K.E.W., C.P.S., J.L., M.K.G., and E.A.S. Investigation: K.E.W., C.P.S., E.B.G., and J.L. Visualization: K.E.W. and C.P.S. Funding acquisition: J.L. and E.A.S. Project administration: J.L. and E.A.S. Supervision: J.L., A.S.A., and E.A.S. Writing – original draft: K.E.W., C.P.S., and J.L. Writing – review and editing: K.E.W., C.P.S., J.L., M.K.G., K.H.G., E.B.G., A.S.A., and E.A.S.

## Competing interests

The authors declare that they have no competing interests.

## Data and materials availability

Data are freely available from the CMU Delphi Research Group to researchers at universities and non-profits at https://cmu-delphi.github.io/delphi-epidata/. All analytic code with dummy datasets is available at https://github.com/HopkinsIDD/inperson-schooling-covid-spring-2021.

## Supplementary Materials

Supplementary Methods

Figs. S1 to S9

Tables S1 to S9

Movie S1

## Notes

### Competing Interest Statement

The authors have declared no competing interest.

### Author Declarations

This study involves only openly available human data, which can be obtained from: https://cmu-delphi.github.io/delphi-epidata/.

