## Supplementary Materials for "In-person schooling and associated COVID-19 risk in the United States over Spring Semester 2021"

**This PDF file includes:**

Supplementary Methods
Figs. S1 to S9
Tables S1 to S9
Caption for Movie S1
References (7,19,32,33)

**Other Supplementary Material for this manuscript include:**

Movie S1 (movieS1.mov)

### Supplementary Methods

Many of the core methods and approach used in this study were represented in Lessler *et al.* (*7*); we reprise them here.

*The United States COVID-19 Trends and Impact Survey:*
The Delphi Group at Carnegie Mellon University and University of Maryland Social Data Science Center COVID-19 Trends and Impact Surveys, in partnership with Facebook (US CTIS) is a cross-sectional survey. Every day, selected Facebook users get an invitation to participate in the survey at the top of their Facebook News Feed. The survey asks questions related to health symptoms, testing, schooling, mental health, vaccination, and preventive behaviors relevant to the ongoing COVID-19 pandemic. It was developed by public health and survey experts and was reviewed and approved by the Institutional Review Boards of the University of Maryland (1587016-3) and Carnegie Mellon University (STUDY2020_00000162). Participants are asked to read a brief text explaining the purpose of the survey and how data are used prior to participating. The survey uses stratified random sampling within US states to provide adequate geographic coverage. The sampling frame is the Facebook Active User Base (FAUB) aged 18+ living in the United States.

To account for systematic demographic differences between the Facebook user sampling frame and the US population, as well as bias related to non-response and coverage, Facebook employs a two-stage weighting process (*19*). In the first stage, inverse propensity score weighting is used to adjust for non-response bias and make the sample more representative of the FAUB. The covariates used in this step are obtained from internal Facebook data, including self-reported age, gender, geographic variables, and other Facebook user characteristics that have been found to correlate well with survey responses in the past. In the second stage, post-stratification is used to balance state-level age and gender distribution among the Facebook population based on the Current Population Survey 2018 March Supplement. The resulting weights are provided as part of the microdata and can be used to adjust estimates to be representative of the US population. In other words, estimates are adjusted for differences between the US population and US Facebook users as well as for the propensity of a Facebook user to take the survey.

Each time Facebook links a user to the survey instrument, it generates a unique, non-informative random ID number and sends it back to CMU. If the user completes the survey (i.e., provides valid answers to a given basic set of survey questions), CMU sends the corresponding ID number back to Facebook and in return receives the survey weight described above. Facebook has no access to the individual survey responses that weight the data, and CMU receives no information linking survey respondents to their Facebook information (*19*, *32*).

To date, eleven waves of US CTIS have been conducted. Each wave includes changes to the survey, such as the addition, deletion, or modification of questions (*33*). Our period of study encompasses Waves 7-8 and 10-11. Wave 7 took place from January 12 to February 7, 2021. Wave 8 was deployed on February 8, 2021, Wave 10 was deployed on March 2, 2021, and

Wave 11 was deployed on May 11, 2021. Wave 9 was skipped and thus not included here. We include data up to June 12, 2021, which is the final full week of data that includes responses about in-person schooling from the Spring Semester 2021. The full text of each round of US CTIS is available at cmu-delphi.github.io/delphi-epidata/symptom-survey/coding.html.

*County-Level Covariates:*
Various county-level measures of socioeconomic status were included in adjusted analyses in addition to county-week-level attack rates. These variables were obtained from the 2014-2018 American Community Survey using the tidycensus package. The county-level variables included in these analyses are (parentheses show variable names in regression equations):

- Total population (**CountyPop**)
- Percent of population that is white (**PctWhite**)
- Percent of households with income level under the appropriate poverty threshold (**CountyPoverty**)
- GINI index of income inequality (**CountyGINI**)
- County type (metropolitan, metropolitan-adjacent, or non-metropolitan, by county size) (**CountyMetroClass**)

*Outcomes and Covariates:*
We considered multiple COVID-19-related outcomes. The three primary outcomes, as reported and experienced by the survey respondent, were:

- COVID-19 like illness (CLI), defined as a fever of at least 100°F, along with cough, shortness of breath, or difficulty breathing in the past 24 hours. [Survey question B2]
- Experience of loss of taste or smell within the last 24 hours. [B2]
- Report of a positive COVID-19 test within the past 14 days. [B10]

Secondary outcomes that addressed household-level risk and motivation for testing, as reported by the survey respondent, included:

- Any member in the household experiencing COVID-19 like illness, defined as fever of at least 100 °F, along with cough, shortness of breath, or difficulty breathing, in the past 24 hours [A1]
- Direct contact in the last 24 hours by the survey respondent with a household member who recently tested positive for COVID-19. A "direct contact" was defined as a conversation lasting more than 5 minutes with someone closer than 6 feet away or physical contact. [C11/C12]
- Report of a positive COVID-19 test result for survey respondent within the past 14 days when testing was indicated (due to either illness or contact with someone who was ill or tested positive for COVID-19) [A10]
- Report of a positive COVID-19 test result for survey respondent within the past 14 days when testing was not indicated (testing was required by an employer, school, or while receiving other medical care and the individual was not ill and had not been in contact with someone who was ill or tested positive for COVID-19). [A10]

We examined the adjusted associations between COVID-19-related outcomes and living with a child in in-person schooling, as well as reported school-based mitigation measures. Responses for in-person schooling practices were provided at the household level (e.g., “Is any child in the household going to in-person classes?”). Respondents were additionally asked whether each of the following mitigation measures was in place where their child(ren) attended in-person classes. The options are stated in the survey exactly as listed below, with the prompt “E3. Do any of the following measures apply to children in your household when they attend in-person classes (pre-K–grade 12)? Please select all that apply” (parentheses show variable names in regression equations):

- Mandatory mask-wearing for students (**StudentMasking**)
- Mandatory mask-wearing for teachers (**TeacherMasking**)
- Student is with the same teacher all day (**SameTeacher**)
- Student is with the same students all day (**SameStudents**)
- Some or all outdoor instruction (**OutdoorInstr**)
- Restricted entry into school (e.g. no parents or caregivers) (**RestrictedEntry**)
- Reduced class sizes (**RedClassSize**)
- Closed cafeteria (**ClosedCafeteria**)
- Closed playground (**ClosedPlayground**)
- Use of separators or “desk shields” in classrooms (**DeskShields**)
- Extra space between desks in classrooms (**ExtraSpace**)
- No school-based extracurricular activities (e.g. sports, clubs, after school care) (**NoExtracurricular**)
- No sharing of books and/or supplies (e.g. each student has their own set at their desk) (**NoShare**)
- Daily symptom screening for those going onto campus (**SymptomScreen**)

We used various other individual- and household-level variables collected through the US CTIS survey in adjusted analyses. Several variables were re-coded to account for out-of-range values, nonsensical responses, and to aid in interpretability. The survey variables used in these analyses were (parentheses show variable names in regression equations):

- **State and zip code of residence.** County of residence is derived from reported zip code; both state and county are used to match to county-level covariates. That is, if the reported county was not within the reported state, county-level covariates were marked as missing.
- **Age of survey respondent,** categorized as 18-24, 25-34, 35-44, 45-54, 55-64, or 65 + years of age (**AgeCategory**) [D2]
- **Gender,** re-coded to whether the survey respondent identified as male or not due to small numbers of non-binary or other respondents (**IsMale**) [D1]
- **Primary occupation** of the survey respondent in the last 4 weeks, where occupations reported by less than 10,000 people (50,000 in the teacher sub-analysis) were collapsed into the “Other” category. (**Occupation**) [Q64]
- **Highest degree** or school level completed by respondent (**EducationLevel**) [D8].
- Indicator for whether the respondent’s performed **work outside of the home for pay** in the last four weeks. [D10]

- **Number of kids, adults, and people 65 years or older in the household.** Negative, non-integer and extreme values were recorded as missing. The number of kids re-coded into eight categories (0,1,2,...,6,7 + ), and the number of adults, people over 65 years, and total household members were re-coded into 11 categories (0,1,2,...,10 + ). Variables for number of known sick contacts, number of people encountered in recent social gatherings, and the number encountered during shopping were not considered due to high levels of missingness. (**NumKidsCategory**, **HHSizeCategory**) [A5]
- **Grade indicators** for whether there is one or more child(ren) in the household in pre-kindergarten or kindergarten; in grades 1 - 5 (elementary school, classically); in grades 6 to 8 (middle school); or in grades 9 - 12 (high school). Only households with at least one school-aged child are included in analyses of household risk. [E1]
- **Full-time and part-time classes indicator.** If at least one school-aged child in the household is in in-person schooling, indicated above, whether or not child(ren) are going to in-person classes full-time and/or part-time. (**FullTime**, **PartTime**). [E2]
- **Masking level,** indicating self-reported frequency of mask or facial covering use in public in the past 5 days (Wave 7) or past 7 days (Waves 8,10-11). Responses were categorized as never, rarely, sometimes, mostly, or always wearing a mask in public, or no reported time spent in public in the past 5 or 7 days (**MaskLevel**). [C14]
- Indicator for whether the survey respondent reported any **out-of-state travel** in the past 5 days (Waves 7-8) or past 7 days (Waves 10-11) (**TravelOutsideState**). [C6]
- Indicators for whether survey respondent reported various **activities** in the past 24 hours, including going to work or school outside place of residence; going to a market, grocery store, or pharmacy; going to a bar, restaurant, or cafe; going to an event with more than 10 people; spending time with someone not currently staying with the survey respondent; and using public transit. In Waves 7-8 this question referred to any activity, while in Waves 10-11 this referred to specifically indoor activity. (**BarRestaurantActivity**, **LargeEventActivity**, **PublicTransitActivity**). [C13]
- **COVID-19 vaccination indicator** for whether or not the respondent had had a COVID-19 vaccination. Responses included ‘Yes’, ‘No’, and ‘I don’t know’; ‘I don’t know’ was re-coded to be missing. (**AnyVaccine**). [V1]
- **Number of COVID-19 vaccine doses received.** If the response was ‘Yes’ to COVID-19 vaccination, options included ‘1 vaccination or dose’, ‘2 vaccinations or doses’, and ‘I don’t know’; ‘I don’t know’ was re-coded to be missing. If the response was ‘No’ to COVID-19 vaccination, the number of vaccine doses was re-coded to be ‘0 vaccinations or doses’. (**VaccineDoses**). [V2]
- **COVID-19 testing indicator** for whether the survey respondent received a COVID-19 test in the past 14 days. Missing values were treated as a negative response (i.e., no test received). Only the subset of individuals who received a test in the past 14 days were used in analyses of positive COVID-19 test outcomes among survey respondents (**Outcome**). [A10]

In addition, we assessed the reported risk of COVID-19-related outcomes associated with education professions among all survey respondents (i.e., not restricted to households with school-aged children). In this analysis, individuals who reported their primary occupation as

“Education” were divided into two separate categories: “K-12 educator”, for respondents who reported being a preschool, kindergarten, elementary, middle, or secondary school teacher; and “Other educator”, for those who reported being a postsecondary teacher, a teacher assistant, other teacher or instructor including special education, or librarian.

*Analysis:*
All analyses were conducted using quasibinomial regression accounting for survey weights using the srvyr package (https://cran.r-project.org/package=srvyr) in the R statistical language.

Analyses of the COVID-19 risk associated with in-person schooling were restricted to households with at least one school-aged child. Analyses of the effects of individual school-based mitigation measures were restricted to households with at least one school-aged child engaged in any in-person schooling. Estimates of the risk of in-person schooling and the impact of individual mitigation measures were adjusted by respondent age, gender, occupation, educational level, masking behaviors, out-of-state travel, and whether they reported a visit to a bar/restaurant/cafe, to an event with more than 10 people, or whether they used public transit; by household size and number of children; and county-week 2-week average attack rate in the past 4 weeks, population, percent white population, percent households in poverty, GINI index of income inequality, metropolitan type, and whether or not they had received any COVID-19 vaccine doses. For each analysis the specific regression equations were as follows. All categorical variables were expanded to be factors:

Unadjusted for control measures

$$\begin{aligned}\text{logit}\ \Pr(\text{outcome}) &= \text{FullTime} + \text{PartTime} + \log_2(\text{CountyAttackRate}) + \text{IsMale} + \text{AgeCategory} + \\ &\quad \text{Occupation} + \text{EducationLevel} + \text{HHSizeCategory} + \text{NumKidsCategory} + \text{MaskLevel} \\ &\quad + \text{TravelOutsideState} + \text{BarRestrauntActivity} + \text{LargeEventActivity} + \\ &\quad \text{PublicTransityActivity} + \text{CountyPop} + \text{CountyPctWhite} + \text{CountyGINI} + \\ &\quad \text{CountyPoverty} + \text{CountyMetroClass} + \text{AnyVaccine}\end{aligned}$$

Adjusted for individual control measures

$$\begin{aligned}\text{logit}\ \Pr(\text{outcome}) &= \cdots\text{<baseline variables>}\cdots + \text{StudentMasking} + \text{TeacherMasking} + \\ &\quad \text{RestrictedEntry} + \text{ExtraSpace} + \text{NoShare} + \text{SameStudents} + \text{RedClassSize} + \\ &\quad \text{SymptomScreen} + \text{SameTeacher} + \text{NoExtracurricular} + \text{ClosedCafeteria} + \\ &\quad \text{DeskShields} + \text{ClosedPlayground} + \text{OutdoorInstr}\end{aligned}$$

Association with in-person schooling by number of mitigation measures

$$\begin{aligned}\text{Logit}\ \Pr(\text{outcome}) &= \text{FullTime}_0 + \text{FullTime}_{1\text{-}3} + \text{FullTime}_{4\text{-}6} + \text{FullTime}_{7\text{-}9} + \text{FullTime}_{10+} + \text{PartTime}_0 + \\ &\quad \text{PartTime}_{1\text{-}3} + \text{PartTime}_{4\text{-}6} + \text{PartTime}_{7\text{-}9} + \text{PartTime}_{10+} + \cdots\text{<baseline variables>}\cdots\end{aligned}$$

Where $\mathrm{FullTime_x}$ and $\mathrm{PartTime_x}$ denote a child engaged in full and part time school with $\mathrm{x}$ interventions reported is in the household. Households reporting children in both full- and part-time school were excluded from this analysis.

<u>Variants</u>

Analyses of risk of COVID-19-related outcomes associated with the Alpha/Delta variants included all respondents living with at least one school-age child. Analyses were adjusted for baseline variables (individual and county-level), individual control measures, county-level attack rate, $\mathrm{AnyVaccine}$, $\mathrm{FullTime}$, $\mathrm{PartTime}$, the fitted combined Alpha+Delta variant proportion, and two interaction terms between variant proportion and each of $\mathrm{FullTime}$ and $\mathrm{PartTime}$. A supplemental analysis was carried out also adjusting for county-level cumulative incidence.

<u>K-12 teachers and other professions</u>

Analyses of the risk of COVID-19-related outcomes among educational professionals were conducted among all respondents with non-missing occupation. An interaction term between occupation types, including K-12 educator, and an indicator for any paid work outside the home in the last four weeks was used to evaluate the baseline risk associated with each occupation, with extra-household work, and with extra-household work within each occupation. These models were adjusted with the same variable set included in models of risk of in-person schooling, excluding the number of children in the household due to missingness.

This same approach was used to examine risk of COVID-19-related outcomes among K-12 teachers by vaccination and work outside the home. Here, the dataset was restricted to K-12 teachers and the model was run with an interaction term between vaccination ($\mathrm{AnyVaccine}$) and an indicator for any paid work outside the home and adjusted for the same variables as above.

<u>In-person schooling and number of vaccine doses</u>

The baseline covariates for these analyses included all the individual- and county-level covariates used above, excluding $\mathrm{FullTime}$, $\mathrm{PartTime}$, and $\mathrm{AnyVaccine}$:

$$\begin{aligned}\mathrm{logit}\ \Pr(\mathrm{outcome}) &= \log_2(\mathrm{CountyAttackRate}) + \mathrm{IsMale} + \mathrm{AgeCategory} + \mathrm{Occupation} + \mathrm{EducationLevel} \\ &+ \mathrm{HHSizeCategory} + \mathrm{NumKidsCategory} + \mathrm{MaskLevel} + \mathrm{TravelOutsideState} + \\ &\mathrm{BarRestrauntActivity} + \mathrm{LargeEventActivity} + \mathrm{PublicTransityActivity} + \mathrm{CountyPop} \\ &+ \mathrm{CountyPctWhite} + \mathrm{CountyGINI} + \mathrm{CountyPoverty} + \mathrm{CountyMetroClass}\end{aligned}$$

Risk associated with in-person schooling by number of vaccine doses was then:

$$\begin{aligned}\mathrm{Logit}\ \Pr(\mathrm{outcome}) &= \mathrm{NoInPerson}_{0\ \mathrm{doses}} + \mathrm{NoInPerson}_{1\ \mathrm{dose}} + \mathrm{NoInPerson}_{2\ \mathrm{doses}} + \mathrm{AnyInPerson}_{0\ \mathrm{doses}} + \\ &\mathrm{AnyInPerson}_{1\ \mathrm{dose}} + \mathrm{AnyInPerson}_{2\ \mathrm{doses}} + \cdots < \text{baseline vaccination analysis} \\ &\text{variables} > \cdots\end{aligned}$$

Where $\mathrm{NoInPerson_x}$ and $\mathrm{AnyInPerson_x}$ denote a child engaged in no in-person school or any in-person school (full- or part-time) with $\mathrm{x}$ number of COVID-19 vaccine doses that the respondent reported having received.

Risk stratified by 0, 1, or 2 vaccine doses was:

$$\text{Logit Pr(outcome)} \\ = \text{FullTime} + \text{PartTime} + \cdots \text{<baseline vaccination analysis variables>} \cdots$$

Risk stratified by full- and part-time in-person schooling was:

$$\text{Logit Pr(outcome)} \\ = \text{VaccineDoses} + \cdots \text{<baseline vaccination analysis variables>} \cdots$$

Where $\text{VaccineDoses}$ was a categorical variable of 0, 1, or 2 vaccine doses.

### Supplementary Figures

**Fig. S1. Comparison of county-month vaccination rates between the survey and CDC data.** (**A**) Proportion of survey respondents living with school-aged children that reported being vaccinated compared to all survey respondents. (**B**) Average monthly differences in proportion vaccinated between respondents living with school-aged children and all respondents by county. (**C**) Proportion of respondents living with school-aged children that reported being vaccinated compared to CDC-reported vaccination rates. (**D**) Average monthly differences in proportion vaccinated between survey respondents living with school-aged children and CDC reported vaccination rates by county. "Vaccinated" represents having received at least one COVID-19 vaccine dose. In (**A,C**) each point represents a county-month. Blue lines indicate fit smooth spline relationships, correlations are Pearson correlation coefficients, and slopes come from a simple linear regression. Limited to county-months with at least 5 survey respondents living with school-aged children (**A-D**) and at least 75% complete CDC data (**C-D**).

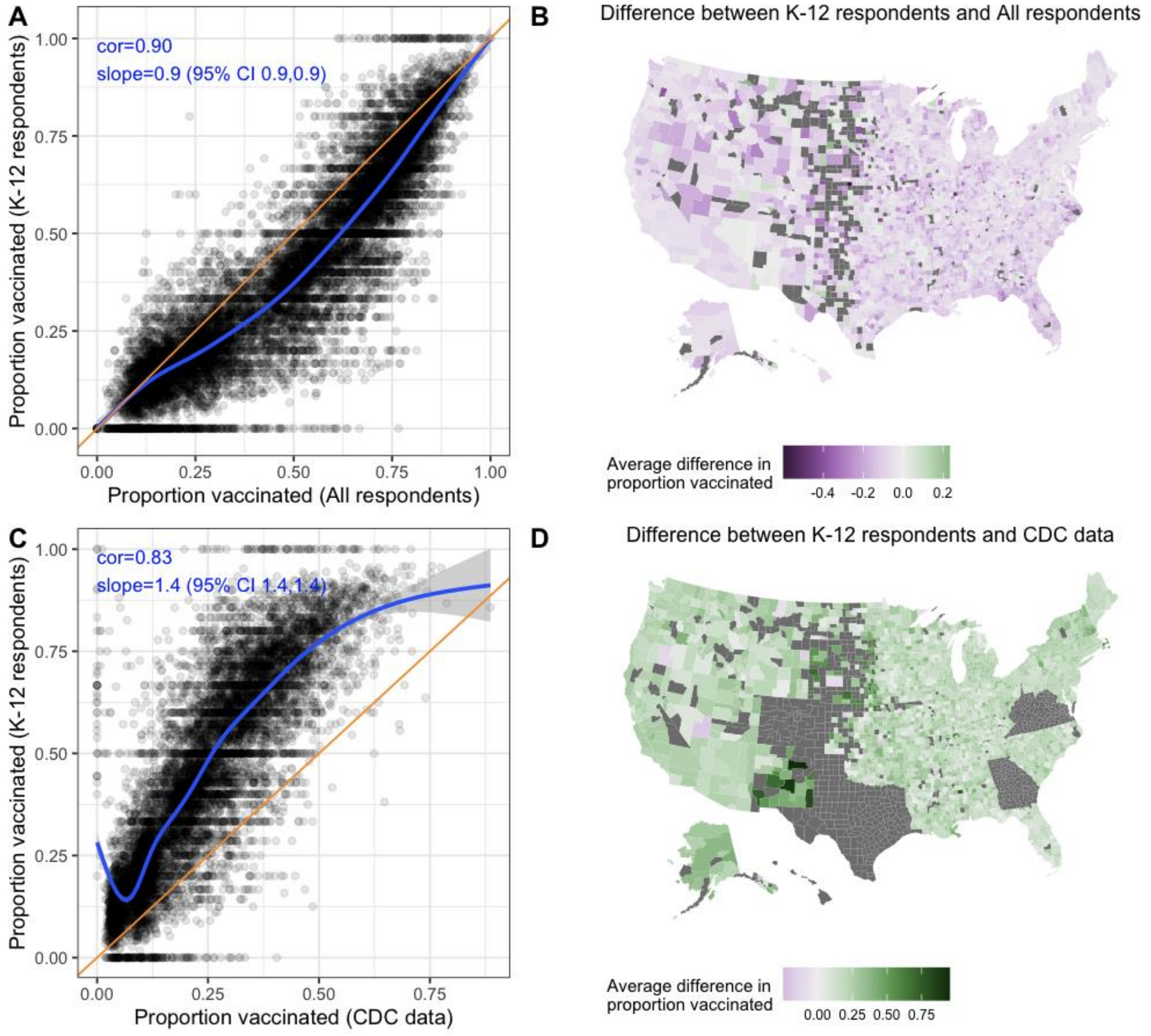

**Fig. S2. Risk associated with in-person schooling by grade level over time.** Odds ratios of COVID-19-related outcomes by full- and part-time in-person schooling compared to no in-person schooling, overall and stratified by grade, adjusted for individual- and county-level covariates. Results are stratified by time periods Jan. 12 to Feb. 28 (Jan-Feb), Mar. 1 to Apr. 30 (Mar-Apr), and May 1 to Jun. 12 (May-Jun). **Note:** grade-stratified analyses include only those respondents who report living with school-aged children in a single grade category, while the overall analyses include all respondents living with school-aged children; thus, the overall estimates do not always fall in-between the grade-specific estimates.

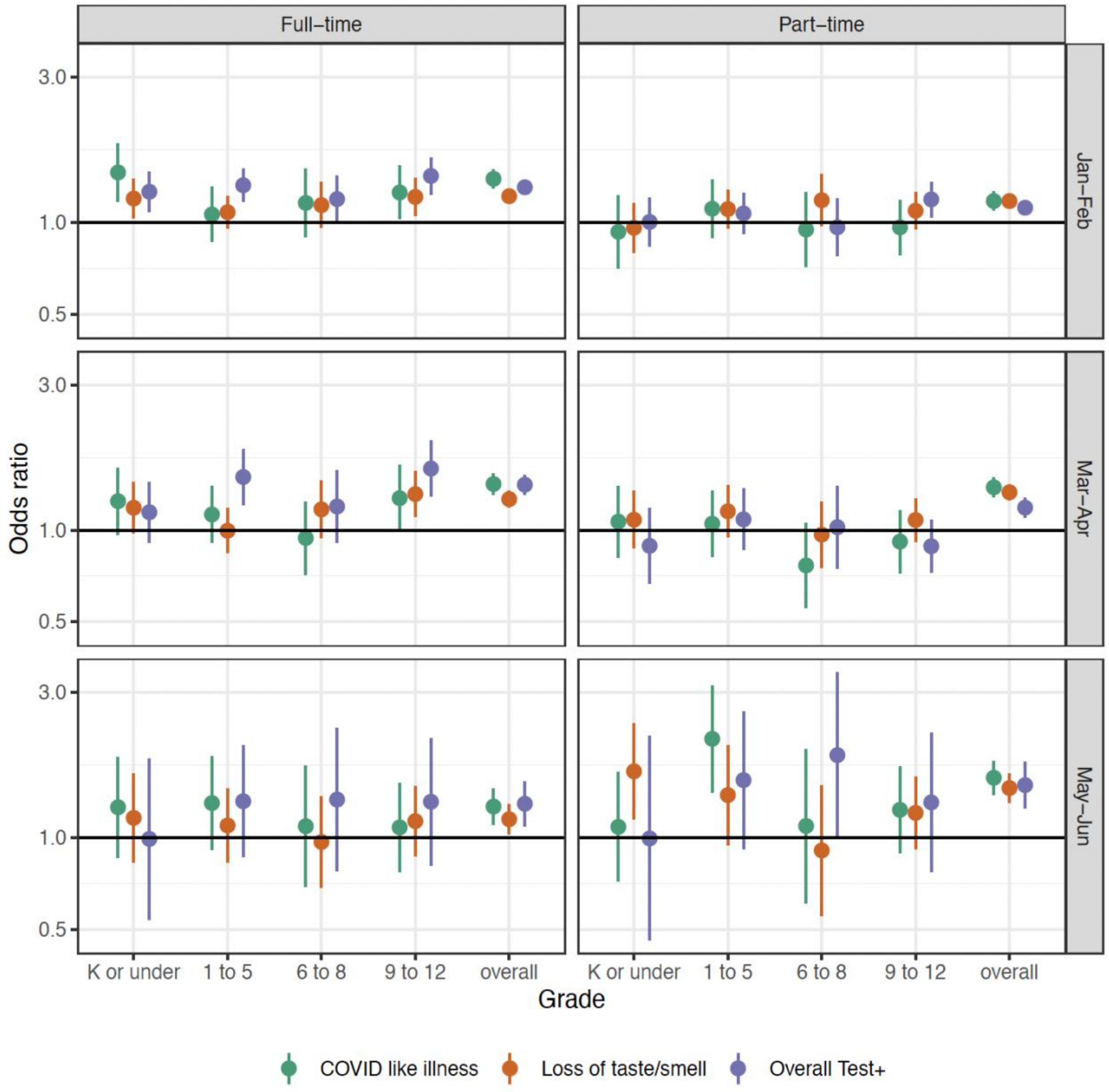

**Fig. S3. Individual mitigation measures over time**. Odds ratio of COVID-19-related outcomes among respondents living with children in in-person schooling who reported each mitigation measure compared to those living with children in in-person schooling who did not report each measure, adjusted for individual- and county-level covariates. Results are stratified by time periods Jan.12 to Feb. 28 (Jan-Feb), Mar. 1 to Apr. 30 (Mar-Apr), and May 1 to Jun. 12 (May-Jun).

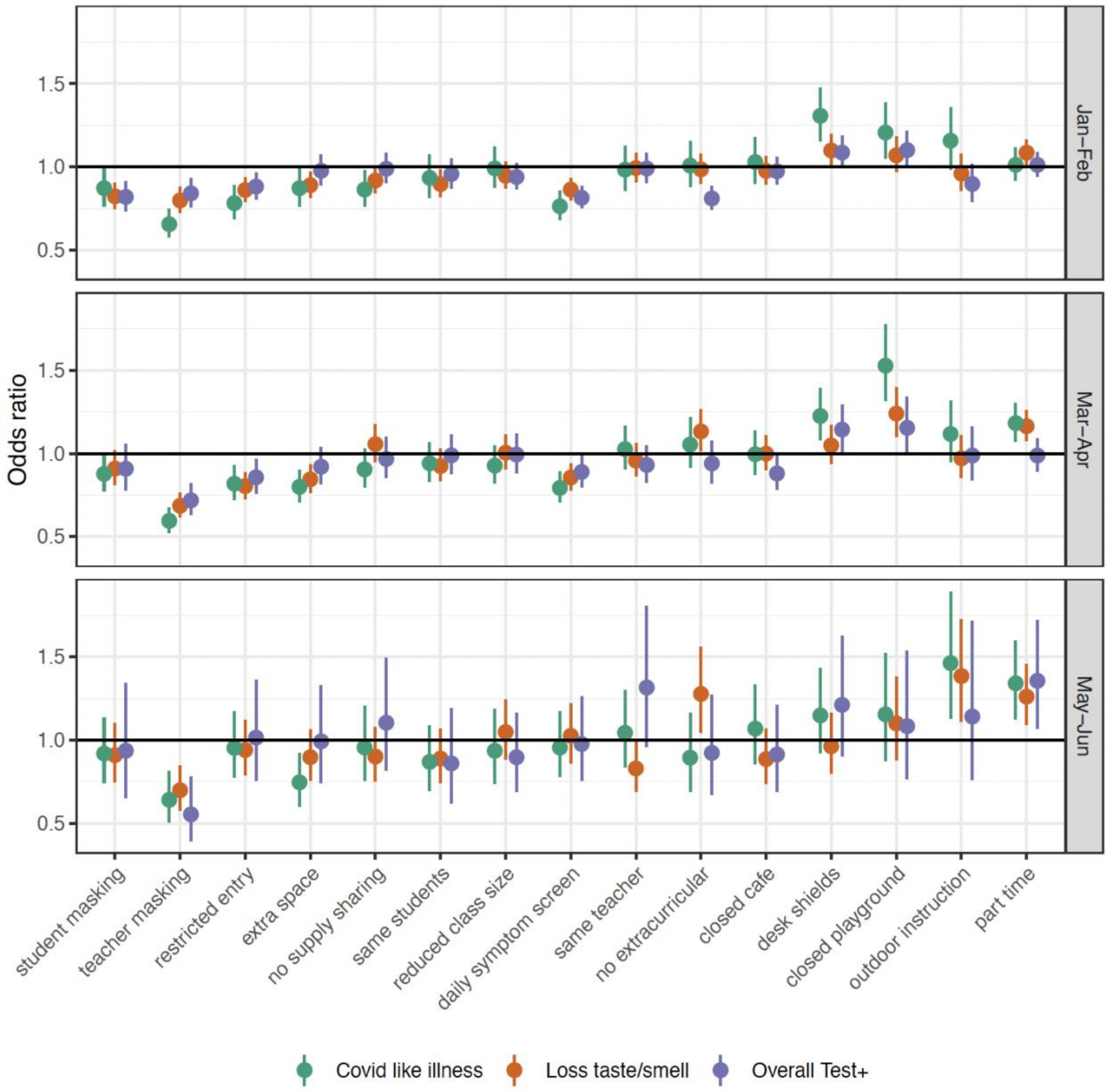

**Fig. S4. Individual mitigation measures and alternative COVID-19-related outcomes**. Odds ratio of alternative COVID-19-related outcomes among respondents living with children in in-person schooling who reported each mitigation measure compared to those living with children in in-person schooling who did not report each measure, adjusted for individual- and county-level covariates across the Spring Semester 2021. Alternative COVID-19-related outcomes include COVID-like illness in any household member (blue), a household member testing positive for SARS-CoV-2 (green), and the survey respondent testing positive when the test was indicated (purple) and when it was not indicated (orange). High uncertainty around non-indicated positive tests reflects a small sample size (533 total non-indicated positive tests).

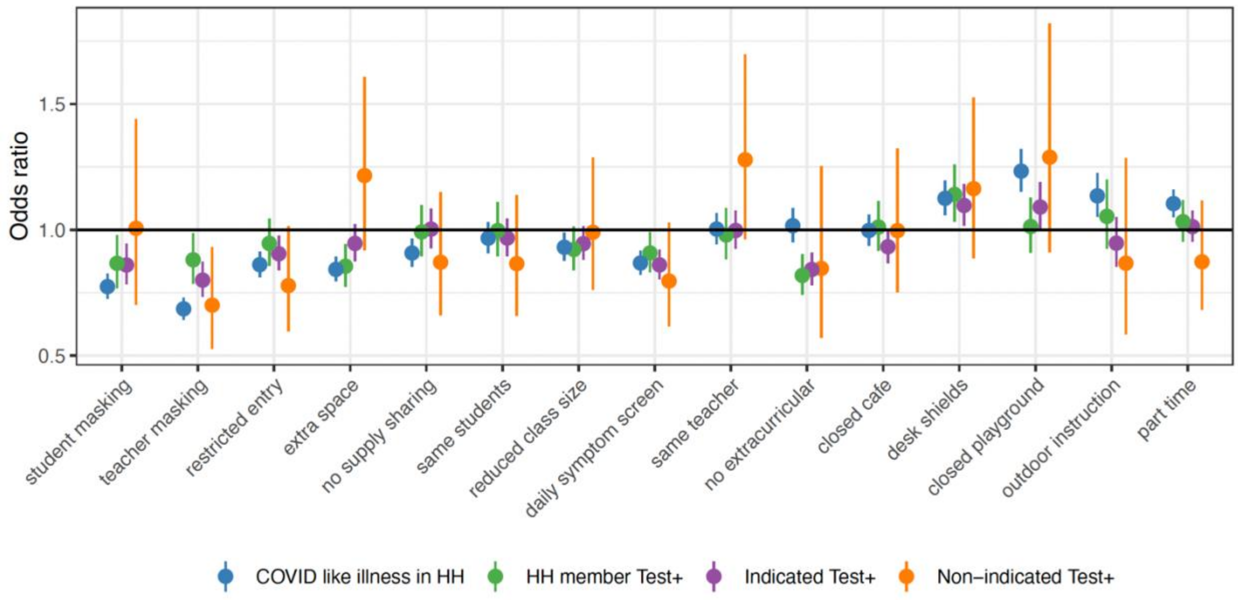

**Fig. S5. Risk associated with in-person schooling by number of reported mitigation measures for alternative COVID-19-related outcomes.** Odds ratio of alternative COVID-19-related outcomes by full- and part-time in-person schooling and number of school-based mitigation measures compared to no in-person schooling, adjusted for individual- and county-level covariates. “Overall” indicates adjusted odds ratios of full-time and part-time schooling with any number of mitigation measures compared to no in-person schooling across the Spring Semester 2021 (Jan 12. – Jun 12.). Alternative COVID-19-related outcomes include COVID-like illness in any household member (blue), a household member testing positive for SARS-CoV-2 (green), and the survey respondent testing positive when the test was indicated (purple) and when it was not indicated (orange). High uncertainty around non-indicated positive tests reflects a small sample size (533 total non-indicated positive tests).

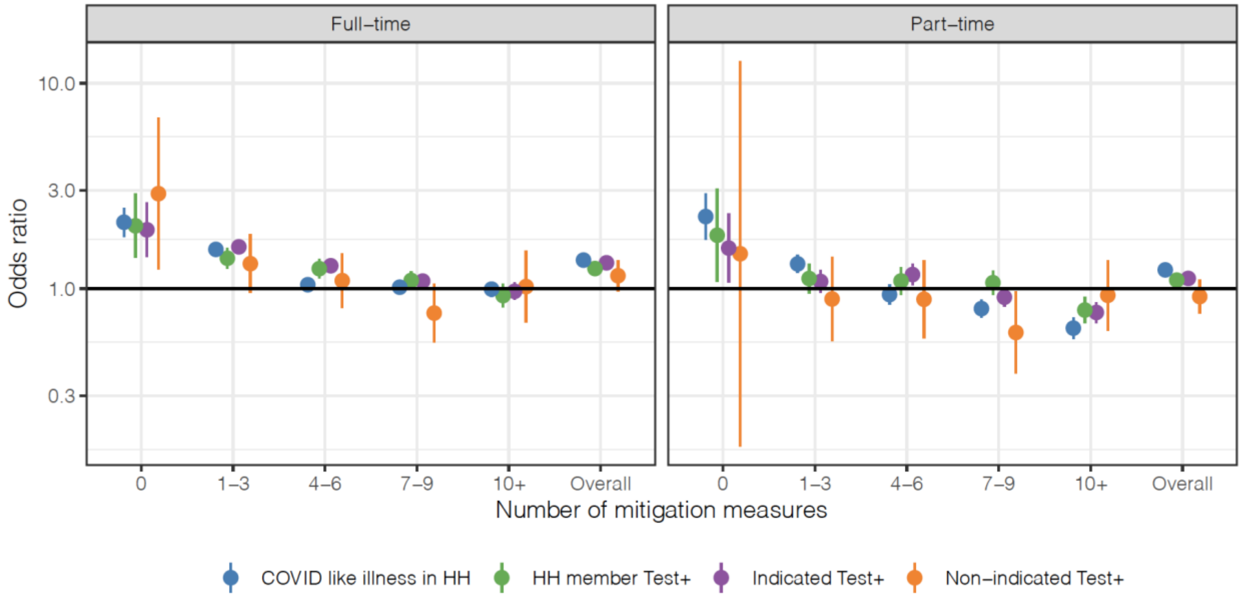

**Fig. S6. Risk associated with in-person schooling and Alpha/Delta variants over time adjusted for cumulative incidence.** Estimated baseline risk (top panel) associated with the proportion of cases due to the Alpha and Delta variants and interaction with part-time (middle panel) and full-time (bottom panel) in-person schooling status. Interaction terms show the additional impact of the variant on top of baseline risk due to part- and full-time in-person schooling. Risk is shown within two-month time strata. Results are adjusted for all baseline covariates as well as cumulative incidence.

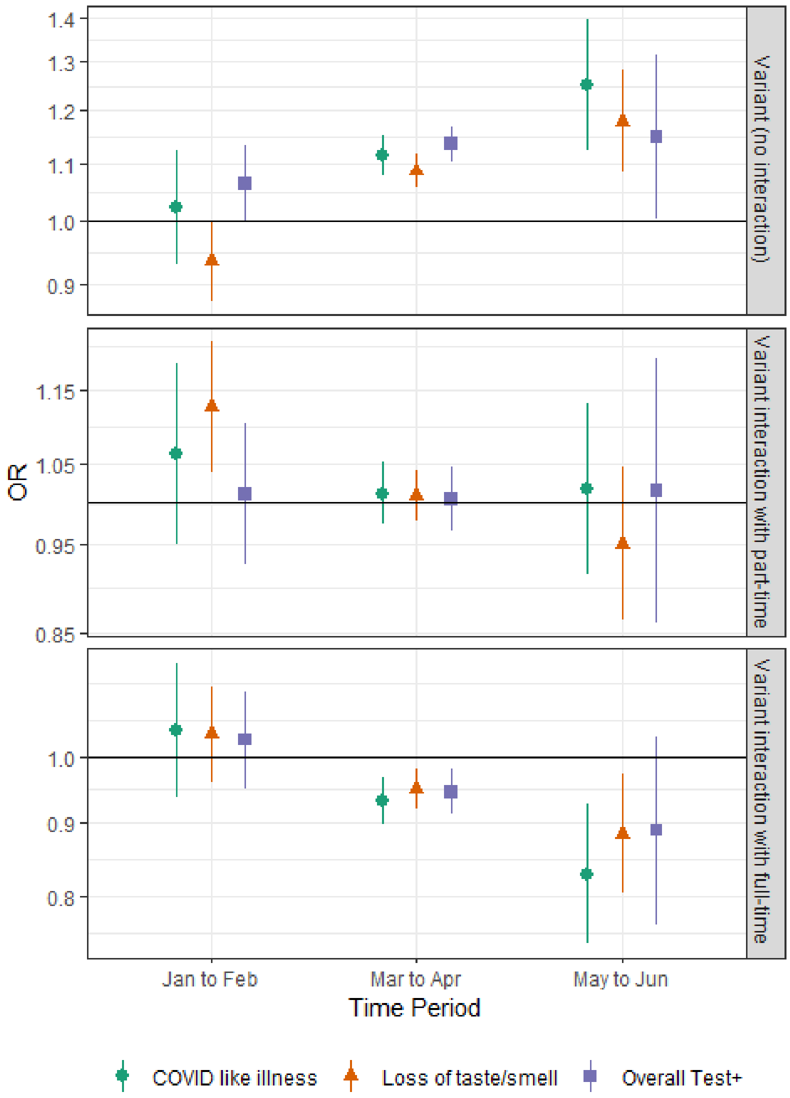

**Fig. S7. Changes in paid work outside the home by occupation.** Percent of all survey respondents in each occupation category that reported working outside of the home for pay in January and June, weighted to account for survey design. Respondents missing occupation designations are excluded. Occupations with fewer than 100,000 respondents are grouped in "Other".

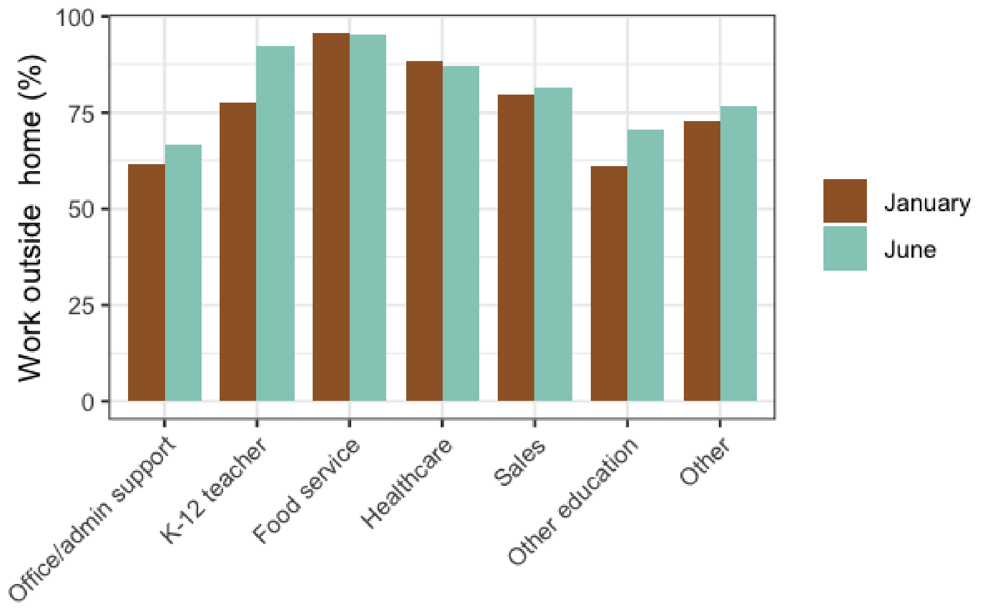

**Fig. S8. Risk by occupation and paid work outside the home over time.** Odds ratio of COVID-19-related outcomes, contrasting office workers not reporting extra-household work for pay to those in other employment categories not reporting work for pay outside the home (top), and to those reporting work for pay outside the home (bottom). The middle row shows the odds ratio (i.e., increased risk) within each category associated with working outside the home compared to no work outside the home. Food service workers are excluded from this analysis because less than 5% reported working exclusively from home. Results are stratified by time periods Jan.12 to Feb. 28 (Jan-Feb), Mar. 1 to Apr. 30 (Mar-Apr), and May 1 to Jun. 12 (May-Jun).

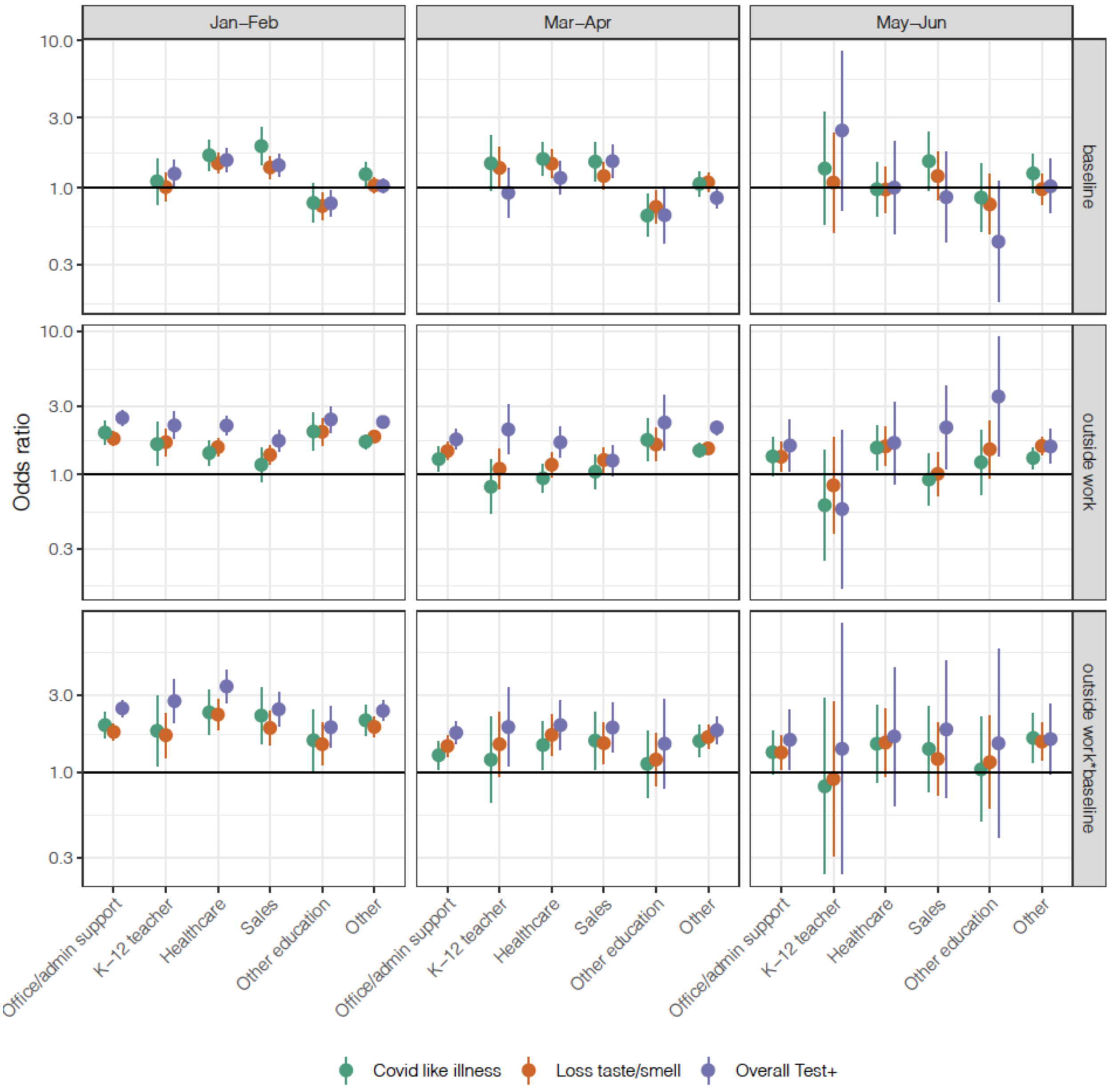

**Fig. S9. COVID-19 risk among K-12 Teachers by vaccination status and paid work outside the home.** Odds ratio of COVID-19-related outcomes, contrasting unvaccinated K-12 Teachers not reporting extra-household work for pay to vaccinated (any doses) K-12 teachers not reporting work for pay outside the home (top), and to those reporting work for pay outside the home (bottom). The middle row shows the odds ratio (i.e., increased risk) within each vaccination category associated with working outside the home compared to no work outside the home.

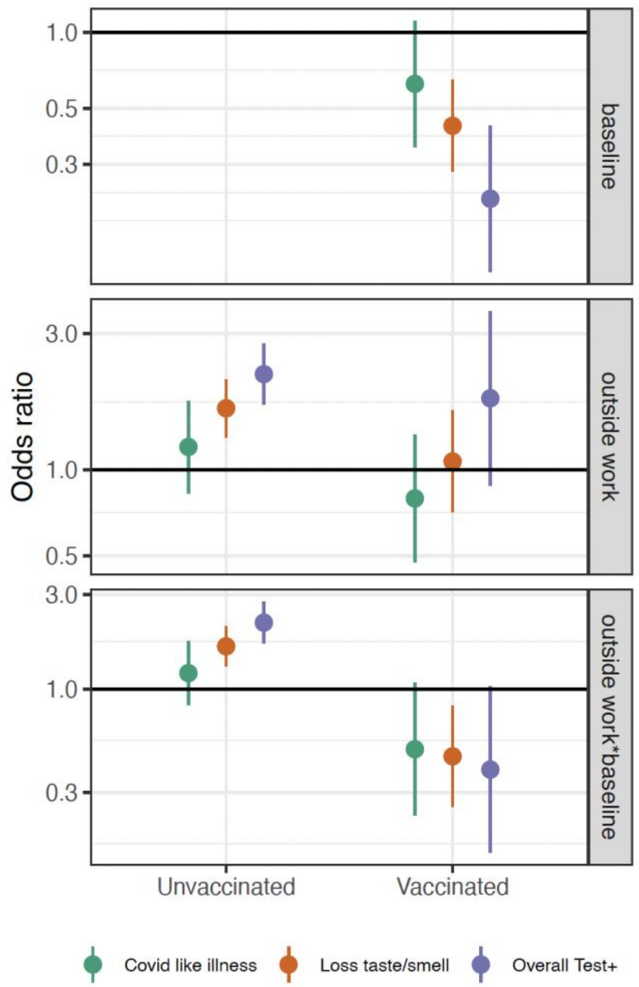

### Supplementary Tables

**Table S1.** Sociodemographic characteristics of participants with ≥1 school-aged child in the household, comparing those reporting no in-person schooling to those reporting any part-time and full-time in-person schooling. Observed and survey-weighted percentages reported.

| | **No child attending in-person schooling** | | **Child attending in-person part-time schooling** | | **Child attending in-person full-time schooling** | |
|---|---|---|---|---|---|---|
| **Variable** | **N (%)** | **Weighted %** | **N (%)** | **Weighted %** | **N (%)** | **Weighted %** |
| *Gender* | | | | | | |
| Female | 306,211 (70%) | 56% | 176,905 (64%) | 51% | 310,845 (66%) | 52% |
| Male | 121,338 (28%) | 40% | 80,083 (29%) | 41% | 132,794 (28%) | 40% |
| Non-binary | 2,655 (0.6%) | 0.80% | 2,128 (0.8%) | 1.10% | 2,893 (0.6%) | 0.90% |
| Other/Prefer not to answer | 7,848 (1.8%) | 2.20% | 7,284 (2.6%) | 3.70% | 11,442 (2.4%) | 3.30% |
| *Age* | | | | | | |
| 18 to 24 | 18,792 (4.3%) | 10% | 11,313 (4.3%) | 9.80% | 14,602 (3.2%) | 7.50% |
| 25 to 34 | 69,380 (16%) | 18% | 39,209 (15%) | 16% | 79,810 (17%) | 19% |
| 35 to 44 | 153,416 (35%) | 32% | 99,304 (37%) | 35% | 187,693 (41%) | 38% |
| 45 to 54 | 116,533 (27%) | 25% | 73,468 (28%) | 26% | 111,962 (25%) | 24% |
| 55 to 65 | 47,251 (11%) | 9.00% | 25,479 (9.6%) | 8.10% | 38,600 (8.5%) | 7.20% |
| 65+ | 30,933 (7.1%) | 5.20% | 16,675 (6.3%) | 5.20% | 23,887 (5.2%) | 4.20% |
| *Child in Pre-K* | | | | | | |
| Yes | 124,994 (28%) | 29% | 89,451 (32%) | 34% | 165,009 (35%) | 36% |
| *Child in Grades 1-5* | | | | | | |
| Yes | 186,138 (42%) | 42% | 131,913 (48%) | 48% | 250,010 (53%) | 53% |
| *Child in Grades 6-8* | | | | | | |
| Yes | 143,361 (33%) | 32% | 107,599 (39%) | 39% | 169,399 (36%) | 36% |
| *Child in Grades 9-12* | | | | | | |
| Yes | 176,544 (40%) | 41% | 129,087 (47%) | 48% | 175,114 (37%) | 38% |
| *Education level* | | | | | | |
| Less than high school | 24,031 (5.6%) | 7.20% | 12,914 (4.9%) | 6.70% | 17,110 (3.8%) | 5.00% |
| High school | 76,881 (18%) | 21% | 40,127 (15%) | 18% | 66,464 (15%) | 17% |
| Some college | 110,466 (26%) | 26% | 60,579 (23%) | 24% | 103,352 (23%) | 24% |
| College/Professional Degree | 157,170 (36%) | 33% | 103,551 (39%) | 36% | 186,148 (41%) | 38% |
| Graduate | 62,740 (15%) | 12% | 45,591 (17%) | 15% | 79,509 (18%) | 16% |
| *Occupation* | | | | | | |
| Office and admin support | 29,118 (6.8%) | 5.80% | 19,620 (7.5%) | 6.40% | 37,389 (8.4%) | 7.20% |
| Arts/entertainment | 7,758 (1.8%) | 1.60% | 4,301 (1.7%) | 1.50% | 6,822 (1.5%) | 1.40% |
| Cleaning and maintenance | 5,086 (1.2%) | 1.50% | 3,054 (1.2%) | 1.50% | 4,924 (1.1%) | 1.40% |

| | | | | | | |
|---|---|---|---|---|---|---|
| Comm/social service | 11,175 (2.6%) | 2.20% | 8,370 (3.2%) | 2.80% | 14,852 (3.3%) | 2.90% |
| Construction and extraction | 3,799 (0.9%) | 1.40% | 3,500 (1.3%) | 2.00% | 6,861 (1.5%) | 2.30% |
| Education | 25,311 (5.9%) | 4.80% | 20,548 (7.9%) | 6.30% | 40,257 (9.0%) | 7.30% |
| Food service | 13,162 (3.1%) | 3.80% | 9,338 (3.6%) | 4.40% | 15,151 (3.4%) | 4.00% |
| Healthcare | 38,036 (8.9%) | 7.80% | 29,678 (11%) | 9.80% | 55,559 (12%) | 11% |
| Installation, maintenance, and repair | 5,651 (1.3%) | 2.00% | 4,370 (1.7%) | 2.50% | 8,078 (1.8%) | 2.70% |
| Not Employed | 182,880 (43%) | 42% | 84,347 (32%) | 32% | 129,134 (29%) | 28% |
| Other | 63,774 (15%) | 16% | 42,250 (16%) | 17% | 73,730 (16%) | 18% |
| Personal care and service | 5,569 (1.3%) | 1.20% | 3,650 (1.4%) | 1.30% | 5,984 (1.3%) | 1.20% |
| Production | 6,090 (1.4%) | 1.70% | 4,486 (1.7%) | 2.10% | 8,537 (1.9%) | 2.30% |
| Protective service | 2,906 (0.7%) | 0.90% | 2,586 (1.0%) | 1.30% | 4,823 (1.1%) | 1.40% |
| Sales | 18,529 (4.3%) | 4.60% | 13,670 (5.3%) | 5.60% | 25,236 (5.6%) | 6.00% |
| Transportation and material moving/delivery | 8,355 (2.0%) | 2.50% | 6,102 (2.3%) | 3.00% | 10,288 (2.3%) | 3.00% |
| *Description of place of residence* | | | | | | |
| Metro - Counties in metro areas of 1 million population or more | 237,835 (54%) | 57% | 128,199 (47%) | 49% | 188,972 (40%) | 42% |
| Metro - Counties in metro areas of 250,000 to 1 million population | 117,666 (27%) | 26% | 75,745 (27%) | 27% | 132,452 (28%) | 27% |
| Metro - Counties in metro areas of fewer than 250,000 population | 37,468 (8.5%) | 7.80% | 30,553 (11%) | 10% | 59,358 (13%) | 12% |
| Nonmetro - Completely rural or less than 2,500 urban population, adjacent to a metro area | 1,884 (0.4%) | 0.40% | 1,690 (0.6%) | 0.60% | 3,562 (0.8%) | 0.70% |
| Nonmetro - Completely rural or less than 2,500 urban population, not adjacent to a metro area | 2,075 (0.5%) | 0.40% | 1,767 (0.6%) | 0.60% | 4,967 (1.1%) | 0.90% |
| Nonmetro - Urban population of 2,500 to 19,999, adjacent to a metro area | 13,488 (3.1%) | 2.80% | 11,203 (4.1%) | 3.90% | 28,352 (6.0%) | 5.90% |

| | | | | | | |
|---|---|---|---|---|---|---|
| Nonmetro - Urban population of 2,500 to 19,999, not adjacent to a metro area | 7,706 (1.8%) | 1.50% | 6,939 (2.5%) | 2.20% | 17,838 (3.8%) | 3.40% |
| Nonmetro - Urban population of 20,000 or more, adjacent to a metro area | 15,462 (3.5%) | 3.20% | 14,399 (5.2%) | 4.90% | 26,297 (5.6%) | 5.40% |
| Nonmetro - Urban population of 20,000 or more, not adjacent to a metro area | 5,938 (1.4%) | 1.20% | 5,108 (1.9%) | 1.70% | 10,337 (2.2%) | 2.00% |
| *Household size (number of persons)* | | | | | | |
| 2 | 24,244 (5.6%) | 4.90% | 11,761 (4.3%) | 3.90% | 21,785 (4.6%) | 4.30% |
| 3 | 99,178 (23%) | 21% | 49,181 (18%) | 17% | 94,005 (20%) | 19% |
| 4 | 135,722 (31%) | 30% | 89,781 (33%) | 31% | 157,389 (34%) | 33% |
| 5 | 81,003 (19%) | 19% | 55,381 (20%) | 21% | 93,530 (20%) | 20% |
| 6 | 45,083 (10%) | 11% | 30,390 (11%) | 12% | 48,898 (10%) | 11% |
| 7 | 21,806 (5.0%) | 5.50% | 14,542 (5.3%) | 5.80% | 22,074 (4.7%) | 5.10% |
| 8 | 11,475 (2.6%) | 3.00% | 7,636 (2.8%) | 3.10% | 11,189 (2.4%) | 2.60% |
| 9 | 5,930 (1.4%) | 1.60% | 3,945 (1.4%) | 1.70% | 5,494 (1.2%) | 1.40% |
| 10+ | 11,176 (2.6%) | 3.00% | 10,471 (3.8%) | 5.00% | 14,247 (3.0%) | 3.90% |

**Table S2.** Monthly COVID-19-related outcomes in different subsets of US CTIS respondents, including total number, observed percent, and percent weighted to account for survey design.

| | COVID-19-like illness | | Loss of taste/smell | | Overall Test+ | |
|---|---|---|---|---|---|---|
| **Month** | **N (%)** | **Weighted %** | **N (%)** | **Weighted %** | **N (%)** | **Weighted %** |
| *All survey respondents* | | | | | | |
| Jan | 908,325 (1.19%) | 1.41% | 908,325 (2.68%) | 2.94% | 908,325 (2.45%) | 2.68% |
| Feb | 1,120,211 (0.95%) | 1.21% | 1,120,211 (2%) | 2.25% | 1,120,211 (1.43%) | 1.64% |
| Mar | 1,181,042 (0.86%) | 1.12% | 1,181,042 (1.62%) | 1.86% | 1,181,042 (0.82%) | 0.97% |
| Apr | 990,232 (0.97%) | 1.24% | 990,232 (1.58%) | 1.82% | 990,232 (0.83%) | 0.95% |
| May | 783,434 (0.94%) | 1.21% | 783,434 (1.52%) | 1.70% | 783,434 (0.39%) | 0.45% |
| Jun | 289,872 (0.97%) | 1.27% | 289,872 (1.54%) | 1.72% | 289,872 (0.03%) | 0.04% |
| *All respondents living with school-aged children* | | | | | | |
| Jan | 210,674 (1.55%) | 1.83% | 210,674 (3.33%) | 3.65% | 210,674 (3.24%) | 3.44% |
| Feb | 245,116 (1.31%) | 1.64% | 245,116 (2.41%) | 2.75% | 245,116 (1.95%) | 2.13% |
| Mar | 246,697 (1.19%) | 1.50% | 246,697 (1.95%) | 2.24% | 246,697 (1.13%) | 1.22% |
| Apr | 209,555 (1.37%) | 1.72% | 209,555 (1.87%) | 2.19% | 209,555 (1.14%) | 1.19% |
| May | 139,515 (1.34%) | 1.76% | 139,515 (1.76%) | 2.11% | 139,515 (0.64%) | 0.67% |
| Jun | 31,216 (1.55%) | 1.99% | 31,216 (1.88%) | 2.17% | 31,216 (0.08%) | 0.09% |
| *Respondents living with school-aged children where no children are engaged in in-person schooling* | | | | | | |
| Jan | 104,528 (1.25%) | 1.36% | 104,528 (3.15%) | 3.38% | 104,528 (3.39%) | 3.68% |
| Feb | 108,911 (0.92%) | 1.01% | 108,911 (2.03%) | 2.18% | 108,911 (1.92%) | 2.13% |
| Mar | 96,937 (0.79%) | 0.90% | 96,937 (1.56%) | 1.67% | 96,937 (1.07%) | 1.16% |
| Apr | 72,783 (0.97%) | 1.02% | 72,783 (1.54%) | 1.58% | 72,783 (1.18%) | 1.18% |
| May | 46,201 (0.86%) | 1% | 46,201 (1.31%) | 1.39% | 46,201 (0.61%) | 0.60% |
| Jun | 10,700 (0.86%) | 0.97% | 10,700 (1.16%) | 1.19% | 10,700 (0.06%) | 0.04% |
| *Respondents living with school-aged children engaged in part-time in-person schooling* | | | | | | |
| Jan | 32,862 (1.06%) | 1.15% | 32,862 (2.57%) | 2.61% | 32,862 (2.57%) | 2.58% |
| Feb | 41,807 (0.81%) | 0.92% | 41,807 (1.88%) | 2.05% | 41,807 (1.57%) | 1.63% |
| Mar | 41,979 (0.68%) | 0.75% | 41,979 (1.48%) | 1.53% | 41,979 (0.94%) | 0.93% |
| Apr | 30,884 (0.77%) | 0.83% | 30,884 (1.2%) | 1.34% | 30,884 (0.81%) | 0.78% |
| May | 19,071 (0.62%) | 0.81% | 19,071 (1.11%) | 1.25% | 19,071 (0.45%) | 0.40% |
| Jun | 3,632 (1.02%) | 1.12% | 3,632 (1.16%) | 1.31% | 3,632 (0.03%) | 0.06% |
| *Respondents living with school-aged children engaged in full-time in-person schooling* | | | | | | |
| Jan | 73,284 (2.21%) | 2.85% | 73,284 (3.92%) | 4.52% | 73,284 (3.32%) | 3.47% |
| Feb | 94,398 (1.98%) | 2.72% | 94,398 (3.09%) | 3.75% | 94,398 (2.16%) | 2.35% |
| Mar | 107,781 (1.75%) | 2.38% | 107,781 (2.49%) | 3.06% | 107,781 (1.25%) | 1.40% |
| Apr | 105,888 (1.83%) | 2.49% | 105,888 (2.29%) | 2.89% | 105,888 (1.2%) | 1.31% |
| May | 74,243 (1.83%) | 2.51% | 74,243 (2.21%) | 2.80% | 74,243 (0.71%) | 0.79% |
| Jun | 16,884 (2.09%) | 2.84% | 16,884 (2.48%) | 2.98% | 16,884 (0.1%) | 0.12% |

**Table S3.** Relative odds of COVID-19-related outcomes by full- and part-time in-person schooling compared to no in-person schooling, either overall among respondents with school-aged children or stratified by grades among households with children in only a single grade.

| | Loss of taste/smell | | COVID like illness | | Test+ | |
|---|---|---|---|---|---|---|
| Grade | adj. OR | 95% CI | adj OR | 95% CI | adj OR | 95% CI |
| January to June | | | | | | |
| Full-time: K or under | 3.30 | 2.90–3.82 | 3.80 | 3.12–4.79 | 3.35 | 2.91–3.92 |
| Full-time: 1 to 5 | 2.89 | 2.62–3.21 | 3.18 | 2.74–3.77 | 4.04 | 3.51–4.72 |
| Full-time: 6 to 8 | 3.10 | 2.70–3.63 | 2.89 | 2.43–3.56 | 3.43 | 2.90–4.15 |
| Full-time: 9 to 12 | 3.51 | 3.11–4.01 | 3.49 | 2.94–4.27 | 4.39 | 3.74–5.25 |
| Full-time: overall | 3.43 | 3.28–3.59 | 3.99 | 3.72–4.30 | 3.84 | 3.64–4.06 |
| Part-time: K or under | 3.01 | 2.62–3.53 | 2.76 | 2.34–3.35 | 2.59 | 2.26–3.03 |
| Part-time: 1 to 5 | 3.19 | 2.81–3.67 | 3.47 | 2.89–4.31 | 2.99 | 2.62–3.46 |
| Part-time: 6 to 8 | 2.92 | 2.51–3.49 | 2.51 | 2.09–3.16 | 2.93 | 2.47–3.57 |
| Part-time: 9 to 12 | 3.05 | 2.75–3.43 | 2.74 | 2.39–3.20 | 2.95 | 2.64–3.34 |
| Part-time: overall | 3.65 | 3.48–3.83 | 3.87 | 3.62–4.14 | 3.24 | 3.08–3.41 |
| January to February | | | | | | |
| Full-time: K or under | 3.32 | 2.81–4.03 | 4.31 | 3.23–6.19 | 3.53 | 2.97–4.33 |
| Full-time: 1 to 5 | 2.95 | 2.60–3.40 | 2.90 | 2.37–3.70 | 3.77 | 3.22–4.50 |
| Full-time: 6 to 8 | 3.13 | 2.61–3.88 | 3.19 | 2.45–4.49 | 3.30 | 2.72–4.15 |
| Full-time: 9 to 12 | 3.37 | 2.86–4.06 | 3.51 | 2.79–4.66 | 4.15 | 3.45–5.13 |
| Full-time: overall | 3.39 | 3.20–3.60 | 4.02 | 3.65–4.47 | 3.69 | 3.46–3.95 |
| Part-time: K or under | 2.61 | 2.22–3.19 | 2.54 | 2.03–3.41 | 2.73 | 2.31–3.35 |
| Part-time: 1 to 5 | 3.03 | 2.60–3.60 | 3.04 | 2.44–4.00 | 2.92 | 2.51–3.49 |
| Part-time: 6 to 8 | 3.27 | 2.65–4.22 | 2.58 | 2.04–3.53 | 2.63 | 2.18–3.31 |
| Part-time: 9 to 12 | 2.99 | 2.59–3.52 | 2.62 | 2.19–3.28 | 3.29 | 2.84–3.91 |
| Part-time: overall | 3.24 | 3.06–3.45 | 3.25 | 2.99–3.54 | 3.06 | 2.89–3.26 |
| March to April | | | | | | |

| | Loss of taste/smell | | COVID like illness | | Test+ | |
|---|---|---|---|---|---|---|
| Grade | adj. OR | 95% CI | adj OR | 95% CI | adj OR | 95% CI |
| Full-time: K or under | 3.27 | 2.66–4.19 | 3.47 | 2.63–4.96 | 3.14 | 2.49–4.21 |
| Full-time: 1 to 5 | 2.71 | 2.32–3.26 | 3.08 | 2.48–4.03 | 4.46 | 3.35–6.34 |
| Full-time: 6 to 8 | 3.22 | 2.56–4.29 | 2.57 | 2.04–3.46 | 3.31 | 2.49–4.80 |
| Full-time: 9 to 12 | 3.72 | 3.03–4.75 | 3.57 | 2.69–5.15 | 4.92 | 3.64–7.14 |
| Full-time: overall | 3.54 | 3.28–3.85 | 4.13 | 3.69–4.66 | 4.10 | 3.70–4.57 |
| Part-time: K or under | 2.95 | 2.39–3.84 | 2.91 | 2.26–4.04 | 2.43 | 1.95–3.25 |
| Part-time: 1 to 5 | 3.17 | 2.58–4.06 | 2.86 | 2.26–3.86 | 2.96 | 2.37–3.93 |
| Part-time: 6 to 8 | 2.63 | 2.12–3.46 | 2.15 | 1.74–2.87 | 2.78 | 2.11–4.05 |
| Part-time: 9 to 12 | 2.94 | 2.50–3.56 | 2.50 | 2.06–3.20 | 2.42 | 2.06–2.95 |
| Part-time: overall | 3.79 | 3.50–4.12 | 3.99 | 3.61–4.44 | 3.27 | 3.00–3.59 |
| May to June | | | | | | |
| Full-time: K or under | 3.20 | 2.30–5.08 | 3.52 | 2.36–6.33 | 2.70 | 1.71–6.19 |
| Full-time: 1 to 5 | 3.00 | 2.29–4.27 | 3.67 | 2.49–6.35 | 3.74 | 2.38–7.44 |
| Full-time: 6 to 8 | 2.64 | 1.98–3.93 | 2.98 | 1.99–5.63 | 3.80 | 2.18–9.90 |
| Full-time: 9 to 12 | 3.11 | 2.38–4.39 | 2.95 | 2.16–4.55 | 3.72 | 2.25–8.34 |
| Full-time: overall | 3.16 | 2.80–3.62 | 3.55 | 3.01–4.28 | 3.65 | 2.98–4.64 |
| Part-time: K or under | 5.22 | 3.15–10.79 | 2.96 | 2.05–5.15 | 2.70 | 1.58–8.62 |
| Part-time: 1 to 5 | 3.98 | 2.58–7.49 | 8.28 | 4.10–23.68 | 4.69 | 2.50–13.51 |
| Part-time: 6 to 8 | 2.48 | 1.74–4.43 | 2.98 | 1.84–7.08 | 6.47 | 2.71–33.17 |
| Part-time: 9 to 12 | 3.34 | 2.51–4.88 | 3.44 | 2.44–5.52 | 3.69 | 2.17–9.11 |
| Part-time: overall | 4.29 | 3.69–5.08 | 4.83 | 4.00–5.98 | 4.43 | 3.48–5.91 |

**Table S4.** Relative odds of COVID-19-related outcomes among respondents living with children in in-person schooling by individual mitigation measures within different time periods.

| | Loss of taste/smell | | COVID like illness | | Test+ | |
|---|---|---|---|---|---|---|
| **Mitigation measure** | **adj. OR** | **95% CI** | **adj OR** | **95% CI** | **adj OR** | **95% CI** |
| January to June | | | | | | |
| student masking | 2.39 | 2.25–2.55 | 2.43 | 2.25–2.63 | 2.38 | 2.21–2.58 |
| teacher masking | 2.08 | 1.98–2.19 | 1.87 | 1.77–1.98 | 2.15 | 2.03–2.29 |
| same teacher | 2.58 | 2.43–2.75 | 2.76 | 2.54–3.03 | 2.71 | 2.53–2.91 |
| same students | 2.48 | 2.34–2.63 | 2.52 | 2.33–2.74 | 2.60 | 2.43–2.80 |
| outdoor instruction | 2.83 | 2.60–3.10 | 3.35 | 2.95–3.86 | 2.60 | 2.38–2.87 |
| restricted entry | 2.36 | 2.24–2.49 | 2.30 | 2.15–2.48 | 2.42 | 2.27–2.58 |
| reduced class size | 2.67 | 2.52–2.85 | 2.57 | 2.38–2.80 | 2.60 | 2.44–2.78 |
| closed cafe | 2.63 | 2.47–2.80 | 2.78 | 2.55–3.05 | 2.54 | 2.39–2.72 |
| closed playground | 3.12 | 2.88–3.41 | 3.73 | 3.29–4.27 | 3.06 | 2.80–3.36 |
| desk shields | 2.87 | 2.69–3.09 | 3.46 | 3.13–3.86 | 3.08 | 2.85–3.35 |
| extra space | 2.39 | 2.27–2.53 | 2.25 | 2.11–2.42 | 2.60 | 2.44–2.80 |
| no extracurricular | 2.94 | 2.74–3.17 | 2.71 | 2.48–3.00 | 2.33 | 2.20–2.49 |
| no supply sharing | 2.63 | 2.48–2.81 | 2.46 | 2.28–2.67 | 2.69 | 2.51–2.90 |
| daily symptom screen | 2.44 | 2.32–2.58 | 2.26 | 2.13–2.41 | 2.35 | 2.23–2.49 |
| part time | 3.17 | 2.99–3.36 | 3.19 | 2.96–3.46 | 2.79 | 2.63–2.96 |
| January to February | | | | | | |
| student masking | 2.28 | 2.11–2.47 | 2.39 | 2.15–2.70 | 2.27 | 2.09–2.49 |
| teacher masking | 2.22 | 2.07–2.41 | 1.93 | 1.78–2.11 | 2.32 | 2.13–2.54 |
| same teacher | 2.70 | 2.48–2.96 | 2.67 | 2.36–3.08 | 2.69 | 2.47–2.96 |
| same students | 2.45 | 2.27–2.67 | 2.54 | 2.25–2.93 | 2.60 | 2.39–2.86 |
| outdoor instruction | 2.61 | 2.35–2.94 | 3.18 | 2.68–3.89 | 2.45 | 2.21–2.76 |
| restricted entry | 2.36 | 2.20–2.56 | 2.19 | 1.99–2.43 | 2.42 | 2.24–2.62 |

| | Loss of taste/smell | | COVID like illness | | Test+ | |
|---|---|---|---|---|---|---|
| Mitigation measure | adj. OR | 95% CI | adj OR | 95% CI | adj OR | 95% CI |
| reduced class size | 2.58 | 2.39–2.80 | 2.69 | 2.40–3.07 | 2.56 | 2.37–2.77 |
| closed cafe | 2.65 | 2.44–2.90 | 2.80 | 2.46–3.24 | 2.65 | 2.44–2.89 |
| closed playground | 2.92 | 2.63–3.26 | 3.34 | 2.86–4.00 | 3.01 | 2.71–3.37 |
| desk shields | 3.00 | 2.74–3.31 | 3.69 | 3.18–4.38 | 2.96 | 2.70–3.27 |
| extra space | 2.44 | 2.26–2.64 | 2.39 | 2.15–2.71 | 2.66 | 2.43–2.92 |
| no extracurricular | 2.68 | 2.46–2.94 | 2.74 | 2.41–3.17 | 2.25 | 2.10–2.42 |
| no supply sharing | 2.50 | 2.32–2.72 | 2.37 | 2.14–2.66 | 2.69 | 2.47–2.95 |
| daily symptom screen | 2.37 | 2.22–2.54 | 2.14 | 1.98–2.35 | 2.26 | 2.12–2.42 |
| part time | 2.96 | 2.76–3.20 | 2.75 | 2.50–3.05 | 2.75 | 2.57–2.96 |
| March to April | | | | | | |
| student masking | 2.48 | 2.25–2.77 | 2.40 | 2.16–2.71 | 2.48 | 2.18–2.88 |
| teacher masking | 1.98 | 1.85–2.15 | 1.81 | 1.69–1.97 | 2.05 | 1.88–2.27 |
| same teacher | 2.60 | 2.37–2.89 | 2.79 | 2.47–3.21 | 2.54 | 2.28–2.86 |
| same students | 2.52 | 2.30–2.79 | 2.56 | 2.29–2.91 | 2.69 | 2.40–3.04 |
| outdoor instruction | 2.64 | 2.34–3.02 | 3.06 | 2.58–3.73 | 2.68 | 2.32–3.19 |
| restricted entry | 2.23 | 2.07–2.43 | 2.27 | 2.06–2.53 | 2.36 | 2.14–2.63 |
| reduced class size | 2.73 | 2.48–3.04 | 2.53 | 2.27–2.85 | 2.70 | 2.41–3.07 |
| closed cafe | 2.72 | 2.46–3.03 | 2.71 | 2.39–3.12 | 2.41 | 2.18–2.69 |
| closed playground | 3.45 | 3.00–4.05 | 4.61 | 3.72–5.91 | 3.17 | 2.70–3.82 |
| desk shields | 2.85 | 2.56–3.22 | 3.40 | 2.94–4.02 | 3.14 | 2.75–3.64 |
| extra space | 2.33 | 2.15–2.54 | 2.22 | 2.03–2.46 | 2.51 | 2.26–2.82 |
| no extracurricular | 3.10 | 2.75–3.55 | 2.87 | 2.49–3.37 | 2.56 | 2.27–2.94 |
| no supply sharing | 2.88 | 2.59–3.23 | 2.47 | 2.22–2.80 | 2.63 | 2.34–3.00 |
| daily symptom screen | 2.35 | 2.18–2.56 | 2.21 | 2.03–2.44 | 2.43 | 2.21–2.70 |
| part time | 3.20 | 2.93–3.53 | 3.26 | 2.92–3.69 | 2.68 | 2.44–2.98 |
| May to June | | | | | | |

| | Loss of taste/smell | | COVID like illness | | Test+ | |
|---|---|---|---|---|---|---|
| **Mitigation measure** | **adj. OR** | **95% CI** | **adj OR** | **95% CI** | **adj OR** | **95% CI** |
| student masking | 2.48 | 2.11–3.01 | 2.51 | 2.10–3.12 | 2.55 | 1.92–3.83 |
| teacher masking | 2.01 | 1.78–2.33 | 1.90 | 1.66–2.25 | 1.74 | 1.48–2.18 |
| same teacher | 2.29 | 2.00–2.70 | 2.84 | 2.31–3.67 | 3.73 | 2.61–6.09 |
| same students | 2.43 | 2.09–2.91 | 2.38 | 2.00–2.97 | 2.36 | 1.86–3.29 |
| outdoor instruction | 3.99 | 3.04–5.61 | 4.32 | 3.10–6.64 | 3.13 | 2.14–5.55 |
| restricted entry | 2.56 | 2.20–3.06 | 2.59 | 2.17–3.22 | 2.76 | 2.13–3.89 |
| reduced class size | 2.85 | 2.42–3.47 | 2.55 | 2.09–3.27 | 2.45 | 2.00–3.20 |
| closed cafe | 2.42 | 2.09–2.90 | 2.91 | 2.36–3.79 | 2.49 | 1.99–3.35 |
| closed playground | 3.01 | 2.41–3.97 | 3.17 | 2.40–4.57 | 2.95 | 2.15–4.64 |
| desk shields | 2.62 | 2.22–3.19 | 3.15 | 2.51–4.19 | 3.36 | 2.47–5.09 |
| extra space | 2.45 | 2.13–2.89 | 2.11 | 1.83–2.52 | 2.70 | 2.10–3.78 |
| no extracurricular | 3.59 | 2.84–4.77 | 2.45 | 1.99–3.20 | 2.52 | 1.95–3.57 |
| no supply sharing | 2.46 | 2.12–2.94 | 2.60 | 2.13–3.33 | 3.02 | 2.26–4.46 |
| daily symptom screen | 2.79 | 2.37–3.39 | 2.60 | 2.18–3.23 | 2.65 | 2.13–3.54 |
| part time | 3.53 | 2.98–4.29 | 3.82 | 3.08–4.95 | 3.88 | 2.91–5.58 |

**Table S5.** Relative odds of COVID-19-related outcomes by full- and part-time in-person schooling and either number of mitigation measures or overall compared to no in-person schooling within different time periods.

| Number of mitigation measures | Loss of taste/smell | | COVID like illness | | Test+ | |
|---|---|---|---|---|---|---|
| | adj. OR | 95% CI | adj OR | 95% CI | adj OR | 95% CI |
| January to June | | | | | | |
| Full-time: 0 | 5.63 | 4.22–7.98 | 5.57 | 4.01–8.37 | 7.96 | 4.82–15.38 |
| Full-time: 1-3 | 4.20 | 3.82–4.64 | 4.59 | 4.02–5.31 | 4.90 | 4.34–5.58 |
| Full-time: 4-6 | 2.52 | 2.37–2.69 | 2.34 | 2.16–2.56 | 3.49 | 3.19–3.85 |
| Full-time: 7-9 | 2.55 | 2.40–2.72 | 2.32 | 2.14–2.53 | 2.83 | 2.63–3.06 |
| Full-time: 10+ | 2.28 | 2.12–2.46 | 2.53 | 2.27–2.87 | 2.65 | 2.43–2.91 |
| Full-time: Overall | 3.30 | 3.16–3.46 | 3.75 | 3.50–4.04 | 3.75 | 3.55–3.97 |
| Part-time: 0 | 5.93 | 3.69–11.29 | 7.46 | 4.05–17.91 | 5.16 | 3.08–10.90 |
| Part-time: 1-3 | 3.44 | 3.05–3.92 | 3.51 | 2.99–4.22 | 3.04 | 2.68–3.50 |
| Part-time: 4-6 | 2.95 | 2.62–3.37 | 2.20 | 1.93–2.57 | 3.15 | 2.78–3.62 |
| Part-time: 7-9 | 2.40 | 2.21–2.63 | 1.93 | 1.76–2.13 | 2.39 | 2.20–2.62 |
| Part-time: 10+ | 2.10 | 1.93–2.31 | 1.84 | 1.68–2.03 | 2.20 | 2.03–2.40 |
| Part-time: Overall | 3.41 | 3.25–3.58 | 3.53 | 3.31–3.77 | 3.09 | 2.94–3.25 |
| January to February | | | | | | |
| Full-time: 0 | 6.92 | 4.47–12.16 | 6.12 | 3.59–13.00 | 6.11 | 3.78–11.77 |
| Full-time: 1-3 | 4.26 | 3.75–4.90 | 5.22 | 4.28–6.53 | 4.36 | 3.77–5.11 |
| Full-time: 4-6 | 2.73 | 2.50–3.01 | 2.47 | 2.18–2.86 | 3.70 | 3.30–4.19 |
| Full-time: 7-9 | 2.54 | 2.33–2.78 | 2.32 | 2.07–2.65 | 2.89 | 2.63–3.20 |
| Full-time: 10+ | 2.25 | 2.05–2.50 | 2.51 | 2.15–3.02 | 2.61 | 2.35–2.94 |
| Full-time: Overall | 3.29 | 3.10–3.49 | 3.75 | 3.41–4.15 | 3.61 | 3.39–3.86 |
| Part-time: 0 | 5.31 | 2.94–13.26 | 5.64 | 2.62–22.40 | 3.66 | 2.08–9.91 |
| Part-time: 1-3 | 3.70 | 3.12–4.50 | 3.46 | 2.77–4.53 | 3.14 | 2.68–3.77 |
| Part-time: 4-6 | 3.05 | 2.63–3.60 | 2.31 | 1.95–2.86 | 3.42 | 2.92–4.11 |

| Number of mitigation measures | Loss of taste/smell | | COVID like illness | | Test+ | |
|---|---|---|---|---|---|---|
| | adj. OR | 95% CI | adj OR | 95% CI | adj OR | 95% CI |
| Part-time: 7-9 | 2.44 | 2.20–2.75 | 2.11 | 1.85–2.49 | 2.60 | 2.33–2.93 |
| Part-time: 10+ | 1.90 | 1.75–2.10 | 1.84 | 1.63–2.12 | 2.11 | 1.92–2.34 |
| Part-time: Overall | 3.14 | 2.96–3.34 | 3.12 | 2.87–3.41 | 2.98 | 2.81–3.18 |
| March to April | | | | | | |
| Full-time: 0 | 5.56 | 3.62–9.84 | 6.65 | 3.86–14.32 | 13.35 | 4.91–68.24 |
| Full-time: 1-3 | 4.26 | 3.65–5.05 | 4.33 | 3.57–5.41 | 5.37 | 4.34–6.86 |
| Full-time: 4-6 | 2.37 | 2.17–2.62 | 2.21 | 1.98–2.51 | 3.33 | 2.85–3.99 |
| Full-time: 7-9 | 2.51 | 2.28–2.79 | 2.31 | 2.05–2.65 | 2.75 | 2.44–3.15 |
| Full-time: 10+ | 2.20 | 1.97–2.51 | 2.47 | 2.11–2.99 | 2.69 | 2.31–3.21 |
| Full-time: Overall | 3.35 | 3.10–3.63 | 3.81 | 3.42–4.29 | 3.99 | 3.60–4.45 |
| Part-time: 0 | 5.86 | 3.03–16.79 | 7.18 | 3.06–32.35 | 8.18 | 3.48–34.67 |
| Part-time: 1-3 | 3.19 | 2.65–3.98 | 3.41 | 2.68–4.60 | 2.81 | 2.30–3.60 |
| Part-time: 4-6 | 2.76 | 2.29–3.49 | 1.97 | 1.68–2.41 | 2.91 | 2.38–3.73 |
| Part-time: 7-9 | 2.27 | 2.01–2.61 | 1.72 | 1.53–1.99 | 2.10 | 1.84–2.45 |
| Part-time: 10+ | 2.30 | 1.96–2.80 | 1.91 | 1.66–2.27 | 2.26 | 1.96–2.69 |
| Part-time: Overall | 3.49 | 3.23–3.79 | 3.60 | 3.27–4.00 | 3.11 | 2.85–3.42 |
| May to June | | | | | | |
| Full-time: 0 | 4.02 | 2.40–9.20 | 3.50 | 2.29–6.62 | 4.61 | 1.88–40.78 |
| Full-time: 1-3 | 3.58 | 2.91–4.58 | 3.90 | 2.97–5.48 | 4.81 | 3.24–8.16 |
| Full-time: 4-6 | 2.26 | 1.96–2.69 | 2.24 | 1.89–2.78 | 2.41 | 1.92–3.27 |
| Full-time: 7-9 | 2.52 | 2.13–3.10 | 2.18 | 1.82–2.76 | 2.27 | 1.82–3.08 |
| Full-time: 10+ | 2.48 | 2.02–3.22 | 2.59 | 1.97–3.81 | 2.29 | 1.73–3.51 |
| Full-time: Overall | 3.06 | 2.71–3.50 | 3.42 | 2.91–4.12 | 3.34 | 2.75–4.21 |
| Part-time: 0 | 6.74 | 2.33–73.68 | 9.51 | 2.98–103.76 | 3.93 | 1.36–421.60 |
| Part-time: 1-3 | 3.13 | 2.35–4.60 | 3.35 | 2.33–5.61 | 3.25 | 1.99–7.51 |

| Number of mitigation measures | Loss of taste/smell | | COVID like illness | | Test+ | |
|---|---|---|---|---|---|---|
| | adj. OR | 95% CI | adj OR | 95% CI | adj OR | 95% CI |
| Part-time: 4-6 | 3.04 | 2.13–5.14 | 2.45 | 1.67–4.81 | 1.91 | 1.41–3.41 |
| Part-time: 7-9 | 2.61 | 1.93–4.05 | 2.00 | 1.57–2.92 | 2.17 | 1.56–3.84 |
| Part-time: 10+ | 2.44 | 1.82–3.76 | 1.64 | 1.33–2.35 | 3.41 | 2.17–6.96 |
| Part-time: Overall | 3.81 | 3.29–4.50 | 4.06 | 3.39–4.99 | 3.85 | 3.03–5.14 |

**Table S6.** Relative odds of COVID-19-related outcomes corresponding to a 10% increase in variant prevalence (A) and interaction between variant prevalence and full- and part-time in-person schooling representing the increase in relative odds among comparted to those without children in part-time (B) and full-time (C) schooling, stratified by time period.

| | **Test+** | | **COVID like illness** | | **Loss of taste/smell** | |
|---|---|---|---|---|---|---|
| **Variant term** | **adj OR** | **95% CI** | **adj OR** | **95% CI** | **adj. OR** | **95% CI** |
| January to June | | | | | | |
| Variant Prevalence (A) | 1.05 | 1.03–1.06 | 1.05 | 1.03–1.07 | 1.02 | 1.01–1.03 |
| Full-time*Variant Interaction (B) | 0.99 | 0.97–1.00 | 0.96 | 0.95–0.98 | 0.99 | 0.97–1.00 |
| Part-time*Variant Interaction (C) | 1.04 | 1.02–1.05 | 1.05 | 1.03–1.07 | 1.05 | 1.03–1.06 |
| January to February | | | | | | |
| Variant Prevalence (A) | 1.03 | 0.97–1.10 | 1.00 | 0.91–1.10 | 0.92 | 0.86–0.98 |
| Full-time*Variant Interaction (B) | 1.04 | 0.96–1.12 | 1.05 | 0.94–1.17 | 1.04 | 0.97–1.13 |
| Part-time*Variant Interaction (C) | 1.01 | 0.92–1.10 | 1.06 | 0.95–1.19 | 1.13 | 1.04–1.22 |
| March to April | | | | | | |
| Variant Prevalence (A) | 1.13 | 1.10–1.17 | 1.11 | 1.08–1.15 | 1.09 | 1.06–1.12 |
| Full-time*Variant Interaction (B) | 0.94 | 0.91–0.98 | 0.93 | 0.90–0.97 | 0.95 | 0.92–0.98 |
| Part-time*Variant Interaction (C) | 1.00 | 0.97–1.05 | 1.01 | 0.97–1.05 | 1.01 | 0.98–1.04 |
| May to June | | | | | | |
| Variant Prevalence (A) | 1.13 | 0.99–1.30 | 1.24 | 1.11–1.38 | 1.18 | 1.08–1.28 |
| Full-time*Variant Interaction (B) | 0.89 | 0.76–1.03 | 0.83 | 0.74–0.93 | 0.88 | 0.80–0.97 |
| Part-time*Variant Interaction (C) | 1.01 | 0.86–1.19 | 1.02 | 0.91–1.13 | 0.95 | 0.86–1.04 |

**Table S7.** Relative odds of COVID-19-related outcomes A) by number of reported vaccine doses and living in a household with a child participating in any or no in-person schooling compared to zero vaccine doses and any in-person schooling, B) with one and two vaccine doses compared to zero doses when data are stratified by none, part-time, and full-time in-person schooling, and C) by part- and full-time in-person schooling compared to no in-person schooling when data are stratified by vaccine doses.

| | Loss of taste/smell | | COVID like illness | | Test+ | |
|---|---|---|---|---|---|---|
| Category | adj. OR | 95% CI | adj OR | 95% CI | adj OR | 95% CI |
| A) OR by vaccine doses and in-person schooling | | | | | | |
| 0 dose & No in-person | 2.49 | 2.40–2.59 | 2.38 | 2.26–2.51 | 2.38 | 2.29–2.47 |
| 1 dose & Any in-person | 1.51 | 1.46–1.57 | 1.67 | 1.57–1.78 | 1.54 | 1.48–1.60 |
| 1 dose & No in-person | 1.39 | 1.35–1.45 | 1.48 | 1.40–1.58 | 1.42 | 1.36–1.48 |
| 2 dose & Any in-person | 1.55 | 1.51–1.60 | 1.91 | 1.82–2.01 | 1.43 | 1.38–1.48 |
| 2 dose & No in-person | 1.29 | 1.25–1.33 | 1.51 | 1.43–1.61 | 1.17 | 1.14–1.20 |
| B) OR by vaccine doses, stratified by in-person schooling | | | | | | |
| Full-time: 1 dose | 1.53 | 1.47–1.60 | 1.72 | 1.61–1.86 | 1.59 | 1.51–1.68 |
| Full-time: 2 dose | 1.58 | 1.53–1.64 | 1.99 | 1.87–2.12 | 1.51 | 1.44–1.60 |
| None: 1 dose | 1.45 | 1.40–1.52 | 1.55 | 1.46–1.67 | 1.50 | 1.43–1.58 |
| None: 2 dose | 1.34 | 1.30–1.40 | 1.61 | 1.50–1.75 | 1.20 | 1.16–1.24 |
| Part-time: 1 dose | 1.49 | 1.39–1.62 | 1.56 | 1.39–1.83 | 1.45 | 1.36–1.58 |
| Part-time: 2 dose | 1.31 | 1.25–1.39 | 1.51 | 1.38–1.69 | 1.17 | 1.13–1.23 |
| C) OR by in-person schooling, stratified by vaccine doses | | | | | | |
| 0 dose: Full-time | 3.36 | 3.20–3.54 | 3.86 | 3.57–4.19 | 3.81 | 3.60–4.05 |
| 0 dose: Part-time | 3.50 | 3.32–3.69 | 3.77 | 3.51–4.06 | 3.01 | 2.86–3.18 |
| 1 dose: Full-time | 3.49 | 2.98–4.20 | 4.44 | 3.44–6.05 | 4.30 | 3.51–5.44 |
| 1 dose: Part-time | 3.39 | 2.88–4.10 | 3.29 | 2.67–4.24 | 3.17 | 2.66–3.89 |
| 2 dose: Full-time | 4.09 | 3.49–4.91 | 4.57 | 3.70–5.85 | 4.03 | 3.19–5.35 |
| 2 dose: Part-time | 4.52 | 3.82–5.46 | 4.06 | 3.40–4.97 | 5.28 | 4.01–7.36 |

**Table S8.** Relative odds of COVID-19-related outcomes among respondents who did, and did not, report any paid work outside the home during different time periods compared to office and administrative staff not working outside of the home for pay.

| | Loss of taste/smell | | COVID like illness | | Test+ | |
|---|---|---|---|---|---|---|
| **Occupation** | **adj. OR** | **95% CI** | **adj OR** | **95% CI** | **adj OR** | **95% CI** |
| January to June | | | | | | |
| Healthcare | 1.35 | 1.19–1.53 | 1.45 | 1.23–1.72 | 1.38 | 1.19–1.59 |
| K12Teacher | 1.05 | 0.87–1.27 | 1.18 | 0.89–1.57 | 1.12 | 0.92–1.36 |
| Other | 1.05 | 0.96–1.14 | 1.17 | 1.04–1.33 | 0.96 | 0.88–1.05 |
| OtherEductation | 0.73 | 0.62–0.85 | 0.73 | 0.59–0.90 | 0.70 | 0.57–0.84 |
| Sales | 1.29 | 1.13–1.47 | 1.66 | 1.37–2.01 | 1.42 | 1.23–1.64 |
| Office and admin support * NA | 1.57 | 1.44–1.72 | 1.51 | 1.33–1.72 | 2.14 | 1.95–2.35 |
| Healthcare * NA | 1.41 | 1.26–1.57 | 1.23 | 1.07–1.41 | 2.00 | 1.76–2.28 |
| K12Teacher * NA | 1.37 | 1.13–1.65 | 1.07 | 0.81–1.42 | 2.04 | 1.67–2.48 |
| Other * NA | 1.69 | 1.60–1.79 | 1.54 | 1.42–1.65 | 2.22 | 2.09–2.37 |
| OtherEductation * NA | 1.81 | 1.54–2.13 | 1.72 | 1.38–2.14 | 2.45 | 2.01–2.99 |
| Sales * NA | 1.26 | 1.12–1.42 | 1.06 | 0.89–1.26 | 1.55 | 1.35–1.77 |
| January to February | | | | | | |
| Healthcare | 1.47 | 1.25–1.72 | 1.67 | 1.31–2.12 | 1.54 | 1.30–1.84 |
| K12Teacher | 1.01 | 0.81–1.26 | 1.11 | 0.78–1.59 | 1.25 | 1.00–1.56 |
| Other | 1.04 | 0.93–1.17 | 1.24 | 1.03–1.48 | 1.03 | 0.92–1.16 |
| OtherEductation | 0.75 | 0.61–0.93 | 0.79 | 0.59–1.07 | 0.79 | 0.64–0.96 |
| Sales | 1.38 | 1.16–1.63 | 1.92 | 1.44–2.56 | 1.43 | 1.19–1.70 |
| Office and admin support * NA | 1.78 | 1.58–2.00 | 1.96 | 1.63–2.37 | 2.49 | 2.21–2.80 |
| Healthcare * NA | 1.55 | 1.35–1.78 | 1.41 | 1.15–1.73 | 2.20 | 1.88–2.57 |
| K12Teacher * NA | 1.68 | 1.35–2.09 | 1.63 | 1.15–2.31 | 2.21 | 1.78–2.73 |
| Other * NA | 1.84 | 1.71–1.98 | 1.70 | 1.52–1.90 | 2.33 | 2.15–2.52 |

| Occupation | Loss of taste/smell adj. OR | 95% CI | COVID like illness adj OR | 95% CI | Test+ adj OR | 95% CI |
|---|---|---|---|---|---|---|
| OtherEductation * NA | 1.99 | 1.60–2.46 | 2.00 | 1.48–2.70 | 2.43 | 1.97–2.98 |
| Sales * NA | 1.37 | 1.17–1.61 | 1.17 | 0.89–1.53 | 1.72 | 1.45–2.04 |
| March to April | | | | | | |
| Healthcare | 1.46 | 1.18–1.81 | 1.57 | 1.21–2.04 | 1.17 | 0.90–1.51 |
| K12Teacher | 1.37 | 0.99–1.89 | 1.47 | 0.95–2.26 | 0.92 | 0.63–1.37 |
| Other | 1.09 | 0.95–1.25 | 1.07 | 0.88–1.28 | 0.86 | 0.73–1.00 |
| OtherEductation | 0.74 | 0.58–0.96 | 0.65 | 0.47–0.90 | 0.65 | 0.42–1.01 |
| Sales | 1.20 | 0.98–1.48 | 1.51 | 1.12–2.02 | 1.52 | 1.18–1.94 |
| Office and admin support * NA | 1.46 | 1.26–1.69 | 1.28 | 1.05–1.57 | 1.76 | 1.50–2.07 |
| Healthcare * NA | 1.17 | 0.96–1.42 | 0.94 | 0.75–1.17 | 1.68 | 1.32–2.14 |
| K12Teacher * NA | 1.09 | 0.79–1.51 | 0.82 | 0.53–1.26 | 2.06 | 1.39–3.07 |
| Other * NA | 1.52 | 1.39–1.66 | 1.47 | 1.31–1.64 | 2.13 | 1.90–2.39 |
| OtherEductation * NA | 1.62 | 1.24–2.11 | 1.74 | 1.24–2.46 | 2.31 | 1.47–3.61 |
| Sales * NA | 1.26 | 1.04–1.53 | 1.05 | 0.79–1.38 | 1.25 | 0.98–1.59 |
| May to June | | | | | | |
| Healthcare | 0.97 | 0.68–1.40 | 0.98 | 0.64–1.49 | 1.00 | 0.49–2.06 |
| K12Teacher | 1.09 | 0.50–2.36 | 1.35 | 0.56–3.26 | 2.45 | 0.71–8.50 |
| Other | 0.98 | 0.78–1.24 | 1.25 | 0.93–1.69 | 1.03 | 0.67–1.56 |
| OtherEductation | 0.77 | 0.49–1.23 | 0.86 | 0.51–1.45 | 0.43 | 0.17–1.10 |
| Sales | 1.20 | 0.82–1.76 | 1.52 | 0.96–2.39 | 0.87 | 0.43–1.75 |
| Office and admin support * NA | 1.33 | 1.05–1.68 | 1.34 | 0.98–1.82 | 1.59 | 1.05–2.42 |
| Healthcare * NA | 1.57 | 1.15–2.15 | 1.54 | 1.08–2.19 | 1.66 | 0.86–3.23 |
| K12Teacher * NA | 0.84 | 0.38–1.83 | 0.61 | 0.25–1.47 | 0.57 | 0.16–2.04 |
| Other * NA | 1.58 | 1.38–1.81 | 1.30 | 1.10–1.54 | 1.57 | 1.19–2.07 |
| OtherEductation * NA | 1.50 | 0.94–2.39 | 1.22 | 0.72–2.05 | 3.51 | 1.35–9.13 |

| | Loss of taste/smell | | COVID like illness | | Test+ | |
|---|---|---|---|---|---|---|
| Occupation | adj. OR | 95% CI | adj OR | 95% CI | adj OR | 95% CI |
| Sales * NA | 1.01 | 0.71–1.43 | 0.92 | 0.61–1.39 | 2.13 | 1.10–4.14 |

**Table S9.** Relative odds of COVID-19-related outcomes among K-12 teachers who were vaccinated with any number of COVID-19 vaccine doses and who did, or did not, report any paid work outside the home during different time periods compared to K-12 teachers who were unvaccinated and not working outside of the home for pay.

| | Loss of taste/smell | | COVID like illness | | Test+ | |
|---|---|---|---|---|---|---|
| **Vaccine status** | **adj. OR** | **95% CI** | **adj OR** | **95% CI** | **adj OR** | **95% CI** |
| January to June | | | | | | |
| vaccinated | 0.43 | 0.28–0.65 | 0.63 | 0.35–1.11 | 0.22 | 0.11–0.43 |
| unvaccinated * NA | 1.64 | 1.30–2.08 | 1.20 | 0.83–1.75 | 2.17 | 1.70–2.77 |
| vaccinated * NA | 1.07 | 0.71–1.61 | 0.79 | 0.47–1.33 | 1.78 | 0.88–3.60 |

**Movie S1. Changes in in-person schooling, vaccination, and variant prevalence. Top left:** County-level smoothed weekly estimates of percent of individuals living with school-aged children participating in part-time or full-time in-person schooling. **Top right:** County-level smoothed weekly estimates of the percent of individuals living with school-aged children that had received any number of COVID-19 vaccine doses. **Bottom left:** State-level smoothed weekly estimates of the proportion of circulating SARS-CoV-2 strains that were the Alpha variant. **Bottom right:** State-level smoothed weekly estimates of the proportion of circulating SARS-CoV-2 strains that were the Delta variant. In-person schooling and vaccination estimated using United States COVID-19 Trends and Impact Survey data; counties with fewer than 20 respondents reporting in-person schooling excluded. Variant prevalence estimated using GISAID sequencing data. Dates represent the last day of each week.
